# Model-based evaluation of alternative reactive class closure strategies against COVID-19

**DOI:** 10.1101/2021.04.18.21255683

**Authors:** Quan-Hui Liu, Juanjuan Zhang, Cheng Peng, Maria Litvinova, Shudong Huang, Piero Poletti, Filippo Trentini, Giorgio Guzzetta, Valentina Marziano, Tao Zhou, Cecile Viboud, Ana I. Bento, Jiancheng Lv, Alessandro Vespignani, Stefano Merler, Hongjie Yu, Marco Ajelli

**Affiliations:** College of Computer Science, Sichuan University, Chengdu, China; School of Public Health, Fudan University, Key Laboratory of Public Health Safety, Ministry of Education, Shanghai, China; Department of Epidemiology and Biostatistics, Indiana University School of Public Health, Bloomington, IN, USA; Center for Health Emergencies, Bruno Kessler Foundation, Trento, Italy; Big Data Research Center, University of Electronic Science and Technology of China, Chengdu, China; Tianfu Complexity Science Research Center, Chengdu, China; Division of International Epidemiology and Population Studies, Fogarty International Center, National Institutes of Health, Bethesda, MD, USA; Laboratory for the Modeling of Biological and Socio-technical Systems, Northeastern University, Boston, MA, USA; ISI Foundation, Turin, Italy; Shanghai Institute of Infectious Disease and Biosecurity, Fudan University, Shanghai, China; Department of Infectious Diseases, Huashan Hospital, Fudan University, Shanghai, China

## Abstract

There are contrasting results concerning the effect of reactive school closure on SARS-CoV-2 transmission. To shed light on this controversy, here we develop a data-driven computational model of SARS-CoV-2 transmission to investigate mechanistically the effect on COVID-19 outbreaks of school closure strategies based on syndromic surveillance and antigen screening of students. We found that by reactively closing classes based on syndromic surveillance, SARS-CoV-2 infections are reduced by no more than 13.1% (95%CI: 8.6%-20.2 %), due to the low probability of timely symptomatic case identification among the young population. We thus investigated an alternative triggering mechanism based on repeated screening of students using antigen tests. Should population-level social distancing measures unrelated to schools enable maintaining the reproduction number (*R*) at 1.3 or lower, an antigen-based screening strategy is estimated to fully prevent COVID-19 outbreaks in the general population. Depending on the contribution of schools to transmission, this strategy can either prevent COVID-19 outbreaks for *R* up to 1.9 or to at least greatly reduce outbreak size in very conservative scenarios about school contribution to transmission. Moving forward, the adoption of antigen-based screenings in schools could be instrumental to limit COVID-19 burden while vaccines continue to roll out through 2021, especially in light of possible continued emergence of SARS-CoV-2 variants.

## Introduction

The novel coronavirus disease 2019 (COVID-19) pandemic caused by severe acute respiratory syndrome coronavirus 2 (SARS-CoV-2) has dramatically changed the life of nearly every human on the planet in 2020. In Europe, Italy was the first country to experience the pandemic and it has been considered a natural experiment for large scale non-pharmaceuticals interventions. The first locally transmitted COVID-19 case in Italy was identified on February 21, 2020; since then, the country went through two distinct epidemic waves. During the first wave, a national lockdown was put in place on March 11, 2020 (1). After the lifting of the lockdown on May 18, 2020 (2), the number of COVID-19 cases remained relatively low throughout the rest of the spring and summer. However, after school reopening and further relaxation of control measures, a second major epidemic wave started in mid-September. At that point, case isolation, contact tracing, and other social distancing measures (e.g., limited size of gatherings, closure of theaters and cinemas (3)) were still in place along with a newly established reactive class closure protocol based on active surveillance of students (4). To counter the rapid rise of cases, a set of nationwide restrictions were imposed by the Italian government on October 14, 2020 (5). New restrictions included an extended mandatory use of face masks, reduction of opening hours or full closure of commercial/recreational venues, and partial or full suspension of in-person education. Control measures gradually increased in the following three weeks (6-8). Since November 6, 2020, more restrictive measures were applied on a regional basis to further mitigate COVID-19 burden (9).

Italy is not an isolated example. A similar upsurge of COVID-19 cases right after school reopening in September-October was observed in several other European countries such as Finland, Ireland, Latvia, Belgium, and Slovakia (10, 11). Moreover, whether associated with school transmission or not, a decrease in the average age of cases was observed, with a larger fraction of cases in the school-age population (12, 13). A similar issue has been experienced in the US in early March 2021 as schools resumed in-person learning amidst rising incidence of the new the B.1.1.7 variant (e.g., Michigan (14)). While the virus is still circulating, and until herd immunity has been reached through natural immunity and vaccination, understanding whether and how in-person education can be maintained is paramount. This is even more important in light of the emergence of new variants with possibly higher transmissibility and/or severity (15-17).

The aim of this paper is twofold. First, we developed a computational model of SARS-CoV-2 transmission to estimate the contribution of the reactive school closure strategies implemented in Italy to mitigate the second major COVID-19 wave and understand the reason of their limited effect. Second, we tested an alternative policy based on rapid antigen-based screening of students. We found that this strategy may have a considerably larger mitigation effect SARS-CoV-2 spread, which may be crucial while COVID-19 vaccines continue to be rolled out throughout 2021.

## Results

### Modeling SARS-CoV-2 transmission

Based on detailed sociodemographic data, we developed a synthetic population of agents representative of the Italian population, whereby each agent in the model corresponds to an individual of the actual population (18). The synthetic population is stratified into three layers representing the network of contacts between (i) household members, (ii) schoolmates and classmates, and (iii) other individuals in the community (which includes both contacts between work colleagues in the workplaces and random encounters in the community). The transmission of SARS-CoV-2 is modeled through the simulation of contacts between agents of the synthetic population in the three layers (see *Materials and Methods* and *SI Appendix* for details). The model allows the explicit simulation of the testing, isolation, and quarantine strategies along with the reactive class closure strategy, as implemented in Italy. In particular, we modeled the following interventions: i) active syndromic surveillance: a symptomatic individual has a probability (50% in the baseline analysis) of being tested through PCR; if positive (i.e., based on a draw from a Bernoulli distribution reflecting the sensitivity of PCR test), the individual is isolated at home for 14 days. Sick isolated individuals can transmit the infection to their household members only; and the members of households are quarantined at home for 14 days as well (95% probability to account for possible sub-optimal adherence); ii) enhanced syndromic surveillance in schools: a symptomatic student has a probability of being tested through PCR (95% in the baseline analysis); iii) if a student is confirmed positive through a PCR test (either by the school symptomatic surveillance system or as an household member of an identified case), the student’s class is closed for 14 days while teaching activities are maintained in the other classes of the same school (see SI Appendix for details and model parameters).

The model leverages data on COVID-19 natural history and SARS-CoV-2 transmission patterns observed in Italy. In particular, we calibrate the model to have a reproduction number (i.e., the mean number of secondary infections caused by a primary infector) of 1.1 when all schools were closed during the summer of 2020 (19) and a household secondary attack rate of 51.5% (20). Then, after schools reopened in mid-September 2020, the reproduction number *R* increased to 1.3-1.9, depending on the Italian region; for instance, over the period October 2-8, 2020 *R* was estimated to be ∼1.3 in Sicily, ∼1.5 in Lazio, ∼1.7 in Veneto (also close to the national average), and ∼1.9 in Lombardy (19). This is similar to the increase estimated for the UK (namely an increase of 0.2-0.7 (21)). To simulate a situation close to that of September/October in Italy, we initialized the simulation with 5% of the population being immune to SARS-CoV-2 (22). As direct quantitative estimates of the contribution of school (or school-related) activities to the increase in the overall transmissibility are unavailable, we consider three scenarios. In the first scenario (F100), we kept the transmission rates in the household and community as estimated for the summer period and set the transmission in the school to obtain the target value of the reproduction number, which corresponds to attributing 100% of the observed increase of the reproduction number in September/October to school transmission. Overall, the total number of infections linked to school transmission in this scenario is 8.4%-16.5% (while 39.3%-41.5% are linked to household transmission and 44.2%-50.1% to community; Fig S2 in *SI Appendix*). The second, more conservative, scenario assumes that 50% of the infections attributed to school transmission in F100 are in fact derived from school and the remainder 50% are due to increased transmission in the community, as other activities involving a predominantly young population had resumed (3). Therefore, in this scenario, the total infections linked to school is estimated to be 4.6%-8.7% (Fig S2 in *SI Appendix)*. Finally, the third scenario (F25) assumes that 25% of the transmission increase was linked to school transmission with the rest occurring in the community (2.5%-5.5% of total infections are linked to school; Fig S2 in *SI Appendix)*. Scenarios F50 and F25 are obtained by increasing the transmission rate in the community and decreasing the transmission rate at school until 50% and 25% of the number of infections generated at school in scenario F100 are linked to school transmission events. Essentially, scenarios F50 and F25 account for the increased number of contacts that inherently occur in the community after school reopening (e.g., increased use of public transport, students’ extracurricular activities). Details are reported in *Materials and Methods* and *SI Appendix* and model parameters are reported in Tab. S1 and S2.

It is important to stress that we do not explicitly model every single measure adopted in Italy to limit transmission in the community (e.g., ban of mass gatherings, closure of cinemas, use of masks) or within school (e.g., desk distancing, mandatory use of masks). These measures are implicit as concerted strategies that result in the different values of *R* (1.3, 1.5, 1.7, and 1.9) – overall transmissibility – and scenarios (F25, F50, and F100) – school relative contribution to transmissibility – explored in this study.

### Impact of the adopted reactive class-closure policy

By forward simulating 1 year of epidemic, we estimate the infection attack rate (which includes all SARS-CoV-2 infections, independently of whether an individual develops symptoms or not) to decrease by less than 15% as compared to a counterfactual scenario with no surveillance in schools, regardless of the school transmission contribution scenario and reproduction number (Fig. 1A). Slightly lower reductions are estimated for the number of COVID-19-related deaths (Fig. 1B); other metrics related to the burden of COVID-19 are reported in *SI Appendix* (Fig. S3). Despite this relatively small mitigation effect, the simulated reactive class closure policy entails a significant cost in terms of missed education: the range of estimates for the mean number of missed school days per student per year is between 22.9 days (95% CI: 19.2-24.0; CI indicates quantile interval in the whole manuscript) and 27.7 days (95% CI: 27.1-28.2) for F25, and between 22.0 days (95% CI: 18.9-23.1) and 25.5 days (95% CI: 24.9-26.1) for F100 (Fig. 1C). This would be comparable to a full school closure for approximately 10% of a school year (i.e., 200 school days). Importantly, this mitigation effect of the strategy is robust to the number of seeds used to initialize the epidemic (Fig. S5 in *SI Appendix*).

**Figure 1.**
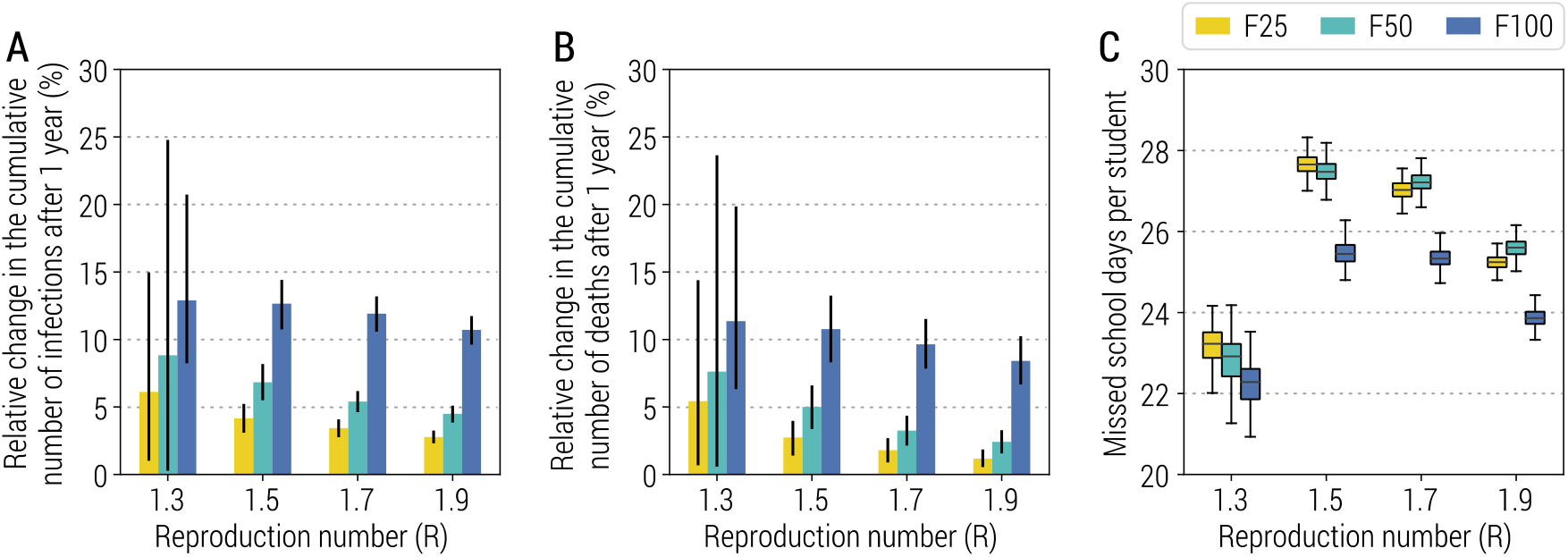
Impact of the reactive class-closure policy based on syndromic surveillance. **A** Relative change in the cumulative number of infections after one year as a function of the reproduction number and for different scenarios about school transmission contribution. The bar corresponds to the mean value, while the vertical line represents 95% quantile intervals; colors refer to the three scenarios F25, F50, F100. Parameters are as the baseline values reported in Tab. S1 and S2. Note that *R* is estimated in the absence of the class-closure strategy. The relative change is defined as the estimated number of infections after 1 year since the introduction of the first infected individual without the implementation of the class-closure strategy minus the one with the class-closure strategy implemented, relative to the estimated number without the implementation of the class-closure strategy. Note that, to exclude spontaneous extinctions from the analysis, only simulations leading to a final infection attack rate of 5% or higher after 1 simulated year are considered. **B** As A, but for the number of deaths. **C** Number of missed school days per student due to the reactive class-closure strategy. In the boxplot, the middle line corresponds to the median, the lower and upper hinges correspond to the first and third quartiles, the upper whisker extends from the hinge to the largest value no further than 1.5IQR from the hinge (where IQR is the inter-quartile range) and the lower whisker extends from the hinge to the smallest value at most 1.5IQR of the hinge. The same definition of the boxplot is used throughout the manuscript. Note that, if no simulations lead to an epidemic with infection attack rate ≥5% for a given combination of model parameters, the number of missed school days is set to 0 by definition.

The implemented reactive class closure strategy is based on the premise of being able to timely identify cases either through symptomatic surveillance of students or through contact tracing. In the baseline scenario, we assumed that the probability of being tested is 95% for symptomatic students, and 50% for symptomatic individuals in the general population, and the time intervals from symptom onset to sample collection and from sample collection to laboratory diagnosis are both 2 days (*SI Appendix*, Tab. S1). To test the timeliness of this strategy, we looked at the number of SARS-CoV-2 infected individuals in an entire school rather than limiting to a class at the time when an infected student is identified, and reactive class closure is triggered. Compared with an average school size of 623.8 students and class size of 23.4 students (23), we estimated that the mean number of infected students in a school at the time when a class is reactively closed ranges from 17.1 (95% CI: 3-46) to 56.7 (95% CI: 4-171), for F50 and *R*=1.3 and 1.9, respectively. This finding is very consistent across different school transmission contribution scenarios (Fig. 2).

**Figure 2.**
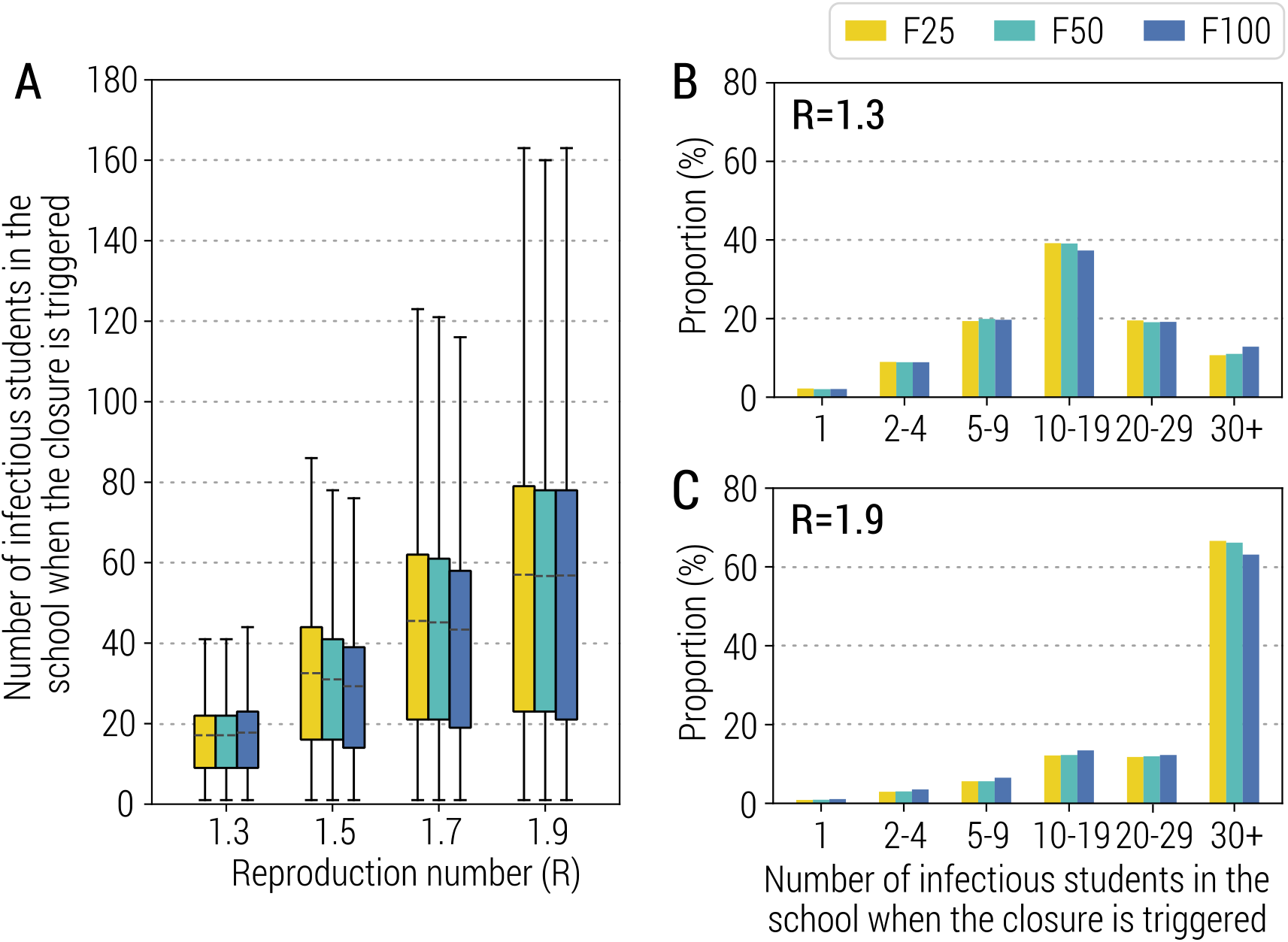
Infectious students at the time of class closure. **A** Number of infectious students in a school for different values of *R* at the time when the class closure is triggered. Parameters are as the baseline values reported in Tab. S1 and S2. Note that *R* is estimated in the absence of the class-closure strategy. Note that, to exclude spontaneous extinctions from the analysis, only simulations leading to a final infection attack rate of 5% or higher after 1 simulated year are considered. **B** Distribution of the number of infectious students in school at any closure for *R=*1.3. **C** As in B, but for *R*=1.9.

To explore whether and to what extent the adopted strategy could be improved or if its limited mitigation benefit is linked to its design, we analyzed alternative scenarios based on an improved testing capacity (in terms of probability of testing symptomatic students or symptomatic individuals in the general population and shorter time intervals from symptom onset to sample collection or laboratory diagnosis). The obtained results are consistent with those obtained for the baseline strategy (Fig. 3 and Fig. S4 in *SI Appendix*), suggesting a structural weakness in the strategy design. These results are also confirmed when homogeneous susceptibility to infection by age is considered and when symptomatic individuals are assumed to be twice more infectious than asymptomatic (Fig. S6 and S7 in *SI Appendix*). Moreover, the results are also consistent in scenarios where the fraction of the initially immune population increases up to 20% (due to either natural infection or vaccination), closer to the to the situation in Italy in early 2021 (24) (Fig. S8 in *SI Appendix*).

**Figure 3.**
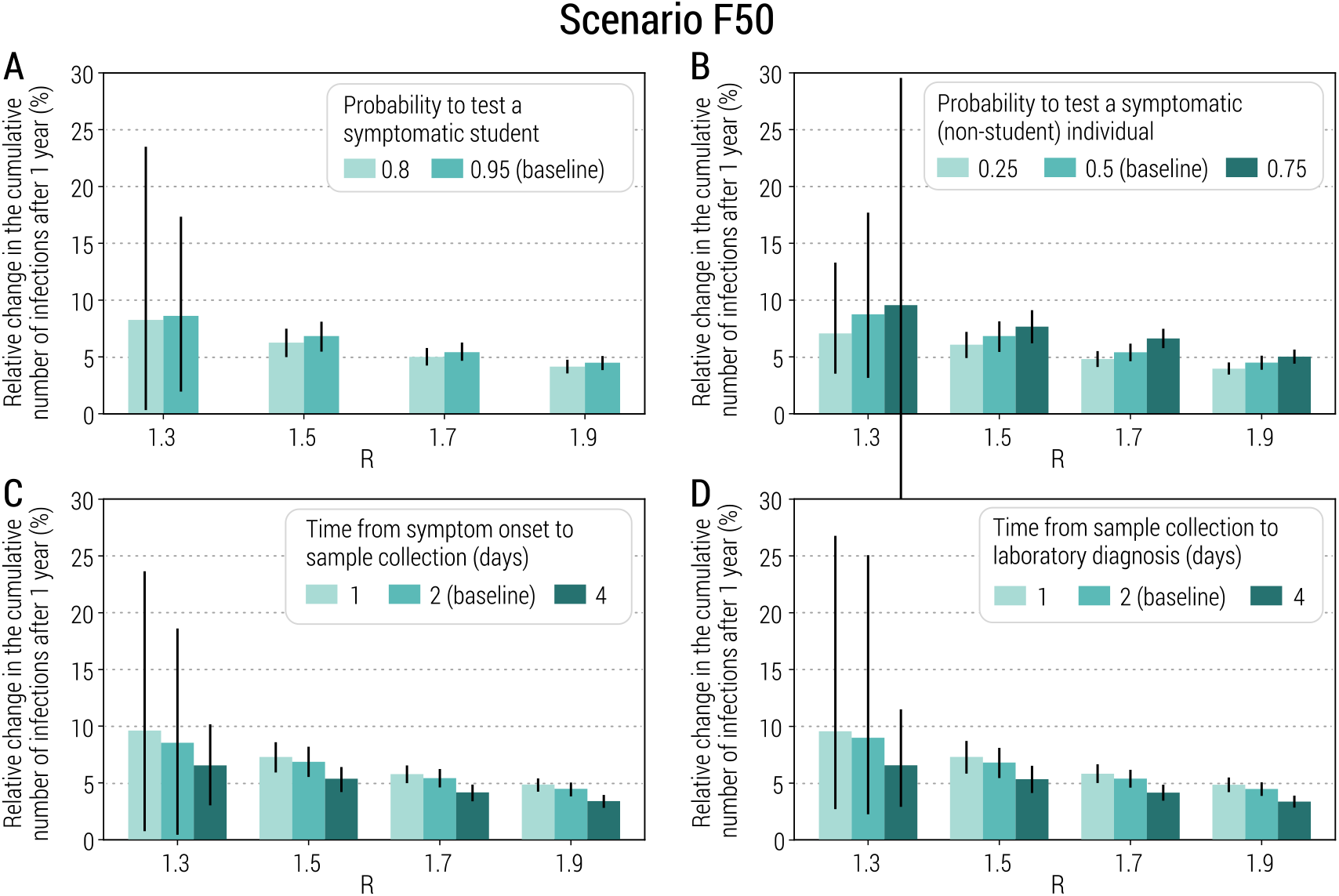
Sensitivity of the class-closure strategy relying on syndromic surveillance to changes in parameters regulating its implementation. **A** Relative change in the cumulative number of infections after one year as a function of the reproduction number and for different values of the probability to test a symptomatic student at school. The bar corresponds to the mean value, while the vertical line represents 95% quantile intervals. Parameters are as the baseline values reported in Tab. S1 and S2. Note that *R* is estimated in the absence of the class-closure strategy and the scenario considered is F50. The relative change is defined as the estimated number of infections after 1 year since the introduction of the first infected individual without the implementation of the class-closure strategy minus the one with the class-closure strategy implemented, relative to the estimated number without the implementation of the class-closure strategy. Note that, to exclude spontaneous extinctions from the analysis, only simulations leading to a final infection attack rate of 5% or higher after 1 simulated year are considered. **B** As A, but for the probability to test a symptomatic (non-student) individual in the community. **C** As A, but for the time from symptom onset to sample collection. **D** As A, but for the time from sample collection to laboratory diagnosis.

### Impact of a reactive school-closure policy

The findings presented thus far suggest that, by the time that classes are reactively closed, outbreaks are already silently taking place in other classes of the same school due to the low probability of young individuals developing symptoms (20), thereby dramatically decreasing the mitigation effect of this strategy. This calls for need to design and implement alternative strategies while vaccines are being rolled out.

Here we tested whether closing the entire school (as compared to the current policy entailing class-based closures) when a SARS-CoV-2 infected student is found represents a more efficient alternative. Our simulation results show that the mitigation effect of this alternative strategy is similar to the current policy, with the consequence of having schools closed for the entire duration of the epidemic (Fig. S9 in *SI Appendix*). This remains true when up to 20% of the population is initially immune (Fig. S10 in *SI Appendix*). Moreover, this strategy fails to timely identify positive students thereby not addressing the main weakness of the baseline scenario.

### Impact of an antigen-based screening strategy

The results presented so far call for the design of a reactive class/school closure policy that goes beyond syndromic surveillance of students. To address this, we consider the potential introduction of screenings of the student population based on antigen tests. This type of testing has the advantages of allowing a quick turnaround time (minutes to hours) and lower costs relative to PCR tests (25). It is worth noting that, in Italy, antigen rapid tests have been used alongside PCR tests to identify SARS-CoV-2 positive individuals since October 30, 2020 (26).

We define an alternative strategy based on repeated screening of students (regardless of symptoms) with rapid antigen testing, while the symptomatic surveillance of the general population remains in place unaltered. We conducted a meta-analysis of the literature to obtain estimates of the sensitivity and specificity of antigen tests, which are 69% (95%CI: 41%-97%) and 99% (95%CI: 97%-100%), respectively (see *SI Appendix* for details). For simplicity, in the model we consider 100% specificity. We tested three different screening schedules: antigen-based tests provided to all students every 3, 7, or 14 days. If a student is found to be SARS-CoV-2 positive either through symptomatic surveillance or antigen screening, the class of such a student is closed for 14 days while the other classes in the school remain open (see *SI Appendix* for details).

We estimate that the strategy based on a weekly antigen screening of students is able to prevent SARS-CoV-2 spread in the population for an *R*=1.3, regardless of the school transmission contribution scenario (Fig. 4A). For scenarios F50 and F100, this strategy is able to reduce the infection attack rate by more than 80% for *R*=1.5 (Fig. 4A) with limited costs in terms of number of missed school days per student (lower than 15.3 days, 95%CI: 2.7-29.7 see Fig. 4B). In a F25 scenario where *R*=1.9, this strategy achieves a 23.8% (95%CI: 23.2%-24.3%) reduction in attack rate (Fig. 4A). Other metrics of COVID-19 burden are reported in Fig. S11 in *SI Appendix*. Under all scenarios considered, the number of missed school days per student remains below 54.7 (95%CI: 52.6-55.9) days. In particular, for *R*=1.7 and F50 (the scenario leading to the highest number of missed school days per student), the mean attack rate reduction per missed school day is 0.77% compared to 0.20% for the syndromic-based screening strategy.

**Figure 4.**
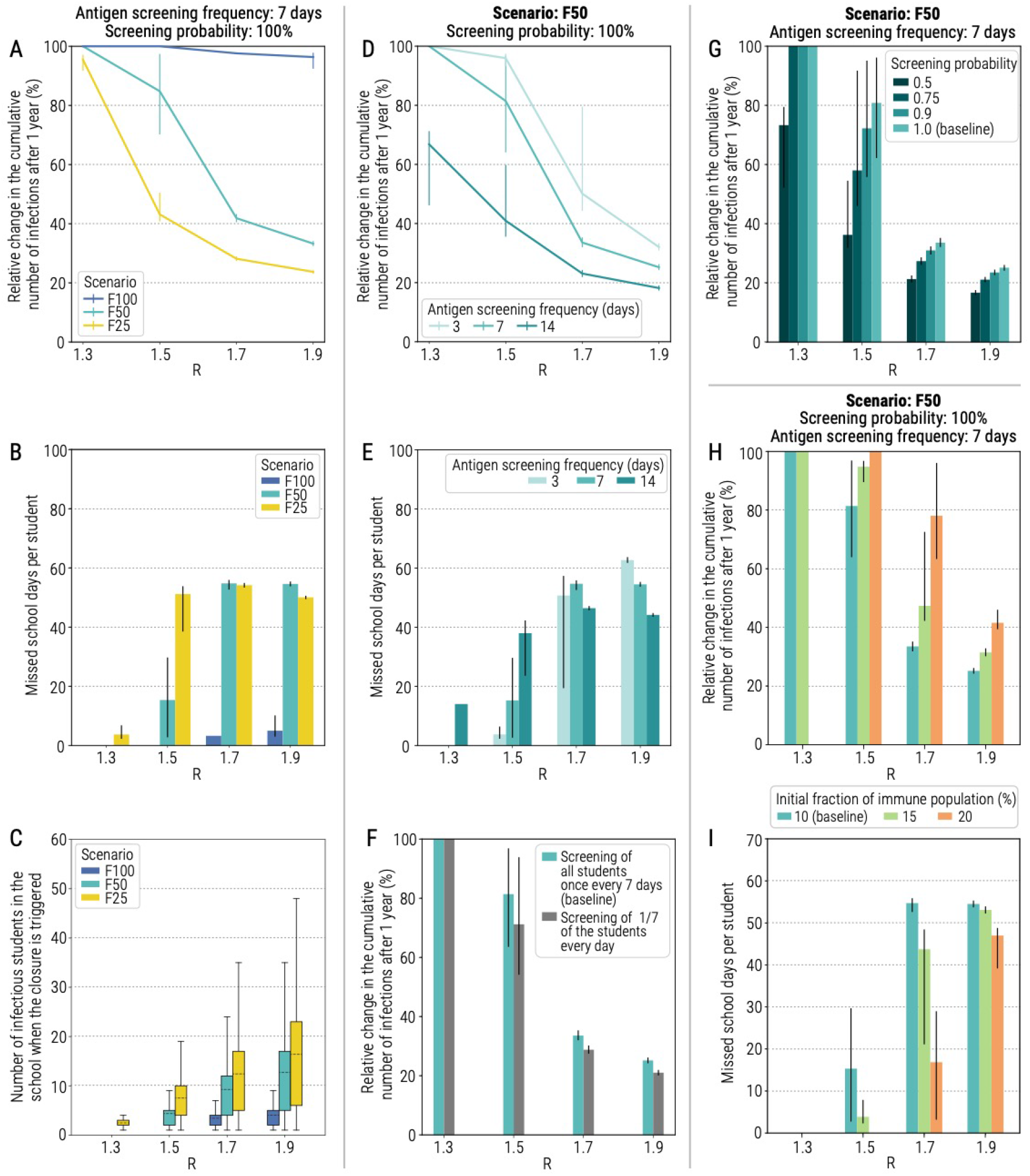
Impact of reactive class-closure policy relying on antigen screening. **A** Relative change in the cumulative number of infections after one year as a function of the reproduction number and for different scenarios about school transmission contribution. The line corresponds to the mean value, while the vertical line represents the 95% quantile intervals; colors refer to the three scenarios F25, F50, F100. The fraction of immune population at the beginning of epidemic is set at 10%, the probability of testing a student at school with the antigen test is 100%, the frequency of the antigen testing is weekly; other parameters are as the baseline values reported in Tab. S1 and S2. Note that *R* is estimated in the absence of the class-closure strategy. The relative change is defined as the estimated number of infections after 1 year since the introduction of the first infected individual without the implementation of the class-closure strategy minus the one with the class-closure strategy implemented, relative to the estimated number without the implementation of the class-closure strategy. Note that, to exclude spontaneous extinctions from the analysis, only simulations leading to a final infection attack rate of 5% or higher after 1 simulated year are considered. **B** Number of missed school days per student due to the reactive class-closure strategy. The bar corresponds to the mean value, while the vertical line represents 95% quantile intervals. Note that, if no simulations lead to an epidemic with infection attack rate ≥5% for a given combination of model parameters, the number of missed school days is set to 0 by definition. **C** Number of infectious students in a school at the time when the class closure is triggered. **D** As A, but for scenario F50 and by varying the frequency of testing (every 3, 7 or 14 days). **E** As B, but for scenario F50 and varying frequency of testing (every 3, 7 or 14 days). **F** Relative change in the cumulative number of infections after one year as a function of the reproduction number when all students are tested in one day, once per week, or when 1/7 of the students at each school are tested every day. **G** As A, but for scenario F50 and by varying the probability of antigen testing (50%, 75%, 90%, and 100%). The bar corresponds to the mean value, while the vertical line represents 95% quantile intervals. **H** As A, but for scenario F50 and by varying the fraction of immune population at the beginning of the simulation (10%, 15%, and 20%). The bar corresponds to the mean value, while the vertical line represents 95% quantile intervals. **I** As B, but for scenario F50 and by varying the fraction of immune population at the beginning of the simulation (10%, 15%, and 20%).

The observed greater mitigation effect of the antigen-based screening strategy as compared to the syndromic-based screening strategy is due to the much better capacity to identify infectious students in a timely manner. For Scenario F50 and a once-a-week testing schedule, the number of infected students in a school at the time when a class is reactively closed never exceeds 4.4 (95%CI: 2-11) students for *R*≤1.5, and 12.7 (95%CI: 2-38) students for *R*=1.9 (Fig. 4C).

By decreasing the testing frequency to once every 2 weeks, we estimate a sharp decrease in the effectiveness of the strategy (Fig. 4D and Fig. S12 in *SI Appendix*), while the number of missed school days per student never exceeds 62.7 days (Fig. 4E). This is likely due to the generation time being lower than 7 days (27-29), suggesting that the testing frequency should be comparable to or shorter than the generation time. We therefore tested the performance of an additional strategy where, instead of testing all students in one single day once per week, we test 1/7 of the student population every day (and thus each student is still tested once a week). Such a strategy would entail a lower number of tests to be performed in a single day, possibly mediating logistical challenges. This alternative analysis shows quantitatively similar results to the baseline analysis (Fig. 4F and Fig. S13 in *SI Appendix*).

We performed sensitivity analyses where we consider a lower coverage of the policy. In particular, we assume 50%, 75%, or 90% of students being tested (rather than 100% considered in the baseline analysis). For scenario F50, changing the coverage of the strategy has little effect on its mitigation power for *R*=1.3, 1.7, and 1.9 (Fig. 4G). In particular, for *R*=1.3 the epidemic is suppressed no matter the coverage of school screening, while for *R*=1.7 or 1.9 school screening substantially reduces attack rates. However, for *R*=1.5, screening coverage becomes a key determinant of the effectiveness of this strategy. With 100% screening coverage this strategy is able to nearly suppress the spread of SARS-CoV-2 (81.2%, 95%CI: 63.1%-96.6% relative change in the cumulative number of infections), while for lower coverages, this strategy mitigates the epidemic rather than suppressing it (Fig. 4G).

With the ongoing transmission and immunization efforts, the population-level immunity in Italy has been growing since the fall of 2020. To understand the contribution of population-level immunity, we performed a sensitivity analysis by increasing the fraction of immune population to 15% and 20% uniformly distributed by age. We estimate that the effectiveness of the strategy remarkably increases with the growing immunity. For instance, if we consider the fraction of immune population to be 20% (likely close to the situation in Italy in early 2021 (22)), the antigen-based reactive school closure strategy is estimated to be capable to successfully prevent outbreaks for an *R* up to 1.5 and F50 (Fig. 4H and Fig. S14 in *SI Appendix*). As expected, the number of missed school days steeply decreases with increasing fraction of immune population (Fig 4I).

## Discussion

Previous studies have investigated the impact of proactive school closure strategies in reducing SARS-CoV-2 transmission (30-39). We provide a quantitative assessment of reactive class closures implemented in Italy since mid-September 2020, in combination with contact tracing and other social distancing measures, to provide a potential explanation of why the adopted strategy was not successful in preventing a second nationwide COVID-19 wave. We propose and evaluate the effectiveness of an alternative strategy that could be applied in the forthcoming months, while vaccines are rolled out. Our modeling results suggest that using syndromic surveillance to trigger case isolation, contact tracing, and the reactive closure of classes with a confirmed COVID-19 infection has a limited impact in mitigating COVID-19 burden (reduction of approximately 10% as compared to no school interventions). In fact, Italy had to rely on a set of restrictions at the regional level and closure of all schools for specific ages as a response to the spread of the second COVID-19 wave (9).

As of March 2021, school-age individuals are not an immediately prioritized group for the ongoing COVID-19 vaccination campaign, reinforcing the need to plan for policies targeting a reduction of their contribution to SARS-CoV-2 transmission. Moreover, it is still unclear when vaccines will be authorized for the vaccination of individuals under 16 years. Our results show that the deployment of antigen tests to perform a routine screening of the student population has the potential to successfully mitigate SARS-CoV-2 spread not only in schools, but in the community at large. This hypothesized policy is estimated to greatly reduce COVID-19 burden as well as fully suppress transmission for a *R* up to 1.3-1.5, depending on the scenario. Multiple reasons contribute to the (estimated) success of this strategy. First, this strategy allows the identification of asymptomatic infections in students – a large fraction of infections in this segment of the population (20).

Second, the identification of asymptomatic students prevents them from transmitting both in the school environment and in the community. Third, the identification of infected students triggers prompt quarantining of their household member, potentially preventing new chains of infections outside the school setting. Finally, the rapid turnaround of antigen-based tests compared to PCR allow for a timely withdrawal of infected individuals from the transmission process.

We note that, in our study, we do not address questions regarding the logistics, feasibility, and acceptability of this strategy. Indeed, an adequate stockpile of antigen-based tests is needed as well as the appropriate logistics surrounding capacity to collect samples from students (e.g., while at school), and compliance to the policy. Moreover, it remains to be determined whether this strategy is cost-effective, accounting for direct and indirect costs associated with the illness, PCR and antigen-based testing, loss of productivity and education. Nonetheless, countries such as the UK are considering such strategies (40). Sporadic attempts have been conducted in specific Italian locations (41) and in several US universities (42, 43). Moreover, in Slovakia a population-wide rapid antigenic screening strategy proved to be feasible and highly effective (44). Compared to the reactive class-closure strategy based on syndromic surveillance, antigen-based screening with 10% of immune population entails greater costs in terms of missed school days for higher *R*, up to 30% of all school days missed per student per year given a maximum *R* of 1.9.

Nonetheless, should the strategy be implemented in mid-2021, the fraction of the immune population will likely be higher than that used in our study, conceivably allowing our proposed strategy to avert new outbreaks entirely, thus bringing educational costs close to zero. Further research is surely needed on to explore this further.

We cannot rule out the possibility that, together with schools reopening and an increase in work and community activities (45), climatic factors may have contributed to the increase of SARS-CoV-2 transmission observed in the fall of 2020 (46). Regardless, the relative contribution of school transmission to the observed trend remains elusive. As such, in our modeling work, we have considered three alternative scenarios on this key parameter. Although the main message of the study remains unchanged, the scenarios highlight noteworthy quantitative differences in the mitigation effect of the proposed policies, calling for further research on this subject.

The developed model is based on a synthetic population of social interactions of the Italian population (47) and on Italy-specific data on COVID-19 epidemiology (infection fatality ratio (48), population immunity (22), hospitalization rates (49), etc.). Nevertheless, the introduced modeling framework is flexible, able to be tailored to other countries to provide insights on the design of COVID-19 control strategies. Moreover, our model could be extended to explicitly simulate additional interventions (such as workplace closure, partial lockdowns, curfews) that can be adopted in conjunction with the proposed strategy, as well as to simulate the parallel roll-out of COVID-19 vaccines and changing of its major target groups.

Moving forward, should logistic challenges be overcome and antigen screening of students gain traction among the public, it could help pursue the “zero-COVID” goal while vaccines are distributed throughout 2021 and beyond. The adoption of this policy could be a game-changing approach, especially as the emergence of new SARS-CoV-2 variants has illustrated the necessity to prepare for the prolonged co-existence with the virus circulation. While the cross-protection from vaccination and natural immunity will hopefully bring the effective reproduction number closer to one (e.g., similar to influenza levels), it will be key to guarantee safe in-person education in the long run.

## Materials and Methods

### Synthetic population

We built a synthetic population of about 0.5 million individuals matching the socio-demographic structure of the Italian population. Each individual of the synthetic population has an associated age, belongs to a certain household, and attends a certain school (and a certain class within the school) if it is a student. School attendance, school sizes, class sizes, and ratio of teacher-to-student are derived from actual data. Details on the construction of the synthetic population are reported in Fumanelli et al. (47). The synthetic population enables us to characterize four different social settings where contacts can occur: home, school, class within school, and the community (which includes any other contact). Given the lack of data about the relative risk of a workplace contact with respect to a community contact, we did not distinguish them in the model and thus a specific layer for workplace contact only was not included. Homogeneous mixing is assumed among individuals in each instance (i.e., a specific household, school, and class) of each social setting. Details are reported in *SI Appendix*.

### SARS-CoV-2 transmission model

We developed an individual-based mechanistic model of SARS-CoV-2 transmission for different social settings to estimate the impact of reactive class closure policies in mitigating COVID-19 burden. Briefly, SARS-CoV-2 transmission is simulated according to an SIR (susceptible, infectious, removed) scheme with pre-symptomatic, symptomatic, and asymptomatic infection compartments. If a susceptible individual *i* is connected with an infectious (either symptomatic, pre-symptomatic or asymptomatic) individual *j*, the susceptible individual can acquire the infection with a setting-specific probability. The model also accounts for age-specific susceptibility to SARS-CoV-2 infection and an infectiousness profile over time that allows matching the empirical distribution of the generation time estimated in the early phase of the epidemic in Italy (50, 51). The incubation period is set to 5 days (52). The probability that an infected individual develops respiratory symptoms and/or fever follows the age-specific ratios estimated for the Italian population (20). Details about the model and its calibration are reported in *SI Appendix*.

## Data Availability

Data referred to in the manuscript can be requested from the corresponding authors.

## Acknowledgments

The authors would like to thank Nicole Samay for her assistance in preparing the figures. Q.-H.L. acknowledge funding from the National Natural Science Foundation of China (No.62003230), Chengdu Science and Technology Bureau (No.2020-YF05-00073-SN), the Fundamental Research Funds for the Central Universities (No.1082204112289), the Science and Technology Department of Sichuan Province (No.2020YFS0009). T.Z. acknowledge funding from the National Natural Science Foundation of China (No.11975071, No.61673085). P.P., F.T., G.G., V.M., and S.M. acknowledge funding from the European Union Grant 874850 MOOD (cataloged as MOOD 000). H.Y. acknowledge funding from the National Science Fund for Distinguished Young Scholars (No. 81525023), Key Emergency Project of Shanghai Science and Technology Committee (No 20411950100). M.L., A.V., and M.A. acknowledge funding from the Cooperative Agreement number NU38OT000297 from the Centers for Disease Control and Prevention (CDC) and the Council of State and Territorial Epidemiologists (CSTE). The study does not necessarily represent the views of CDC and CSTE. The funders had no role in the design and conduct of the study; collection, management, analysis, and interpretation of the data; preparation, review, or approval of the manuscript; and decision to submit the manuscript for publication.

