## Supplementary materials for "Model-based evaluation of alternative reactive class closure strategies against COVID-19"

**This PDF file includes:**

Supplementary text
Figures S1 to S14
Tables S1 to S3
SI References

### Contents

1 SARS-CoV-2 transmission model ... 3
1.1 Synthetic population ... 3
1.2 Transmission model ... 3
1.3 Reproduction number ... 6
1.4 Simulated intervention strategies ... 7
1.5 Model parametrization ... 10
2 Reactive class-closure strategy based on syndromic surveillance ... 12
2.1 Additional results for the baseline analysis ... 12
2.2 Initial number of seeds ... 14
2.3 Homogeneous susceptibility to infection by age ... 15
2.4 Infectiousness of asymptomatic vs. symptomatic individuals ... 16
2.5 Initial fraction of immune population ... 17
3 Reactive school-closure strategy based on syndromic surveillance ... 19
4 Reactive class-closure strategy based on rapid antigen screening ... 21
4.1 Additional results for the baseline analysis ... 21
4.2 Antigen screening frequency ... 22
4.3 Initial fraction of immune population ... 24
5 SI References ... 25

### 1 SARS-CoV-2 transmission model

#### 1.1 Synthetic population

We adopted the algorithm used for modeling contact patterns in European populations (1) to build a synthetic population of about 500,000 individuals. The modelled population mimics the sociodemographic structure of the actual Italian population in terms of age structure, household size distribution and within household age composition, school attendance rates, school size distribution, class size distribution, and the number of teachers per students.

Each single individual is explicitly represented in the model as an agent. The network of contacts between individuals can be described by three different layers accounting for contacts between household members, between schoolmates (and classmates within each school), and all other contacts occurring in the general community (i.e., contacts related to leisure activities, use of transportation means, and contacts occurring among work colleagues, etc.). Households are defined as $n_h$ disconnected components (i.e., the total number of households ) grouping a number of individuals sampled from the actual Italian household size distribution (2). In the model, individuals' age is determined by an algorithm tailored to reproduce realistic age-gaps between household members for a given household size and to match the actual Italian age distribution (1, 2).

Similar to the household layer, schools of different types (primary, middle, and high schools) are defined as $n_s$ (i.e., the total number of schools) disconnected components with size sampled from the Italian distribution of school size. Individuals are assigned to different schools taking into account age-specific school enrollment rates and the teacher-to-student ratio associated to each school type, as reported in the official statistics. Briefly, students are randomly assigned to one single class and one single school using a heuristic approach that allows us to mirror the average, minimum, and maximum class size reported in the official records, and the age composition within classes. Teachers of each school are randomly sampled from the individuals of the synthetic population based on age distribution of teachers in Italy.

The general community layer is represented by one single fully connected component consisting of all individuals in the population and aims at representing the network of all contacts occurring outside households and schools.

#### 1.2 Transmission model

In the synthetic population, SARS-CoV-2 transmission is simulated using a discrete-time stochastic Markov process. We consider that each individual can be characterized by one of five mutually exclusive epidemiological states (corresponding to the states of the Markov process): susceptible (i.e., individuals who may acquire infection after exposure to SARS-CoV-2 infected individuals), infectious pre-symptomatic (i.e., individuals who are not showing any clinical sign or symptom but that are able to transmit the infection and will develop symptoms in the future), infectious symptomatic (i.e., individuals who developed symptoms are able to transmit the infection), infectious asymptomatic (i.e., individuals who are able to transmit the infection, but that will not developed symptoms), and removed (i.e., individuals who recovered from the infection gaining immunity against re-infection).

In the model, SAR-CoV-2 transmission occurs upon contacts between susceptible and infectious individuals taking place in one of the four transmission settings (household *h*, school *s*, class *c* within schools *s*, and the general community *r*). At each time step *t* (corresponding to one day), the probability that a susceptible individual *i* is infected through a contact with an infectious individual *j* in setting $l$ is modeled as:

$$\mathrm{p}_{[\mathrm{j}\rightarrow\mathrm{i}]}(\mathrm{t}) = \beta_l \delta(a_i) \chi(\mathrm{s_j}) \phi(t-\tau_j)/n_{l(j)} \qquad 1.1$$

where:

- $\beta_l$ is the daily per contact transmission rate, shaping the risk of infection due to interactions with an infectious individual ($\mathrm{day}^{-1}$) in setting $l$, $l \in \{h, s, c, r\}$.
- $n_{l(j)}$ is the number individuals in the household ($l = h$), school ($l = s$), class ($l = c$) where individual *j* belongs to. Since all individuals are potentially in contact in the community, for $l$=r, $n_{l(j)}$ corresponds to the total number of individuals in the population.
- $a_i$ is the age of individual *i*.
- $\delta(a_i)$ is relative susceptibility to SARS-CoV-2 infection at age $a_i$ (Tab. S1).
- $\mathrm{s_j}$ is dummy variably identifying whether individual $j$ is symptomatic or asymptomatic. To determine whether the infectious individual of age $a_i$ will develop clinical symptoms or not, we draw a random sample from a Bernoulli distribution with probability $s(a_i)$ at the time when *i* acquires the infection (Tab. S2).
- $\chi(\mathrm{s_j})$ is the relative infectiousness of asymptomatic to symptomatic individuals.
- $\phi(t-\tau_j)$ is the infectiousness of individual $j$ at time $t-\tau_j$, where $\tau_j$ is the time at which individual *j* was infected and *t* is the time step of the simulation (Tab. S1).

As just mentioned, the model explicitly considers infectiousness over time in agreement with empirical epidemiological estimates (3, 4). Essentially, in the first days since the infection, an individual is capable of transmitting the infection, but the probability is extremely low. The infectiousness over time is chosen such that the generation time ($T_g$ - the mean time interval between time of infection of a secondary infectee and the time of infection of its primary infector) in the model mirrors the distribution estimated in (5). The incubation period is not related to the transmission process (which instead depends on the infectiousness over time), but defines the time interval between acquiring the infection and the development of symptoms. In the model, the incubation period is assumed to last 5 days on average (6). The incubation period is key in our analysis, as the syndromic surveillance is based on the detection of symptomatic individuals (either at school or in the community).

According to the estimates reported in previous studies, we assume that the infectiousness of pre-symptomatic, symptomatic, and asymptomatic individuals is the same, $\chi = 1$ (3). However, we performed a sensitivity to explore how model outcomes change when asymptomatic individuals are assumed to be half infectious compared to symptomatic and pre-symptomatic cases (i.e., $\chi=0.5$).

**Table S1**. Summary of model parameters.

| Param | Description | Value (or range) / Procedure | Sensitivity analysis | Reference |
|---|---|---|---|---|
| **Natural history** | | | | |
| - | Incubation period (days) | 5 days | - | Zhang et al. (6) |
| $\mathrm{T_g}$ | Generation time (days) | 6.6 days [95%IQR: 0.7-19.0] | - | Cereda et al. (5) Lavezzo et al. (7) |
| *R* | Reproduction number (when schools are open) | 1.3, 1.5, 1.7, 1.9 | - | ISS (8) |
| - | Initial immunity | 5% | 10%, 15%, 20% | Marziano et al. (9) |
| **Parameters related to the infection transmission process** | | | | |
| $\beta_h$ | Transmission rate in household | Set to obtain a household secondary attack rate of 51.5% according to Poletti et al. (10) | - | Derived |
| $\beta_r$ | Transmission rate in the community | Set to obtain the desired value of *R* when schools are closed | - | Derived |
| $\beta_s$ | Transmission rate between schoolmates | Set to obtain the desired value of *R* when schools are open | - | Derived |
| $\beta_c$ | Transmission rate between classmates | $\beta_c = \beta_s * \frac{N_c}{N_s}$ where $N_c = 6.3$ (mean daily number of contacts by students with classmates) and $N_s = 1.5$ (mean daily number of contacts by students with schoolmates) according to Litvinova et al. (11) | - | Derived |
| $\delta_a$ | Susceptibility to infection by age *a* | $\delta_a = 0.58,\ a < 15$; $\delta_a = 1,\ 15 \leq a < 65$; $\delta_a = 1.65,\ a \geq 65$ | $\delta_a$=1 for all ages | Hu et al. (3) |
| $\chi$ | Transmissibility of asymptomatic relative to symptomatic individuals | 100% | 50% | Hu et al. (3) |
| **Reactive school closure** | | | | |
| | Number of days of class closure | 14 days | - | ISS (12) |
| **PCR test** | | | | |

| | | | | |
|---|---|---|---|---|
| $\alpha_s$ | Probability of being tested if symptomatic for students | 95% | 80% | Assumed |
| $\alpha_p$ | Probability of being tested if symptomatic for non-students (passive surveillance) | 50% | 25%, 75% | Assumed |
| $\phi_p$ | Sensitivity of PCR test | 0.979 | - | Xiao et al. (13) |
| $T_{ST}$ | Delay from symptom onset to sample collection | 2 | 1, 4 | ISS (14) |
| $T_{TR}$ | Delay from sample collection to PCR results | 2 | 1, 4 | ISS (14) |
| **Rapid test** | | | | |
| T | All students are tested every T days | 3, 7, 14 | - | Assumed |
| $\phi_a$ | Sensitivity of antigen test# | 0.69 | - | Meta-analysis of sensitivity (Fig. S1) |
| **Routine quarantine** | | | | |
| q | Probability of quarantine the household if one member tested as positive | 0.95 | - | Assumed |

### The result of the test is available on the same day the test is performed.

**Table S2.** Age-specific parameters regulating COVID-19 disease burden.

| **Age group (years)** | **Probability that an infected individual will develop respiratory symptoms and/or fever** (10) **(mean, 95% CI)** | **Probability that an infected individual will require hospitalization (**15) **(mean, 95% CI)** | **Probability that an infected individual will require ICU treatment** (10) **(mean, 95% CI)** | **Infection fatality risk** (16) **(mean, 95% CI)** |
|---|---|---|---|---|
| **0-14** | 17.3% (13.1-22.3%) | 2.5% (1.0-5.1%) | 0.0% (0.0-1.3%) | 0.0% (0.0-1.3%) |
| **15-19** | 22.2% (8.6-42.3%) | 7.4% (0.9-24.3%) | 0.0% (0.0-12.8%) | 0.0% (0.0-12.8%) |
| **20-39** | 23.7% (20.2-27.6%) | 5.3% (3.5 -7.5%) | 0.4% (0.0-1.4%) | 0.0% (0.0-0.7%) |
| **40-59** | 30.9% (28.1-33.9%) | 13.0% (11.0-15.2%) | 0.9% (0.4-1.7%) | 0.3% (0.1-0.9%) |
| **60-69** | 31.8% (27.7-36.1%) | 17.2% (14.0-20.8%) | 2.6% (1.4-4.5%) | 1.4% (0.6-2.9%) |
| **70-79** | 41.5% (36.2-47.0%) | 27.5% (22.8-32.6%) | 7.2% (4.6-10.5%) | 6.9% (4.4-10.1%) |
| **≥ 80** | 63.9% (55.9-71.4%) | 43.0% (35.2-51.1%) | 18.4% (12.7-25.3%) | 18.4% (12.7-25.3%) |

##### 1.3 Reproduction number

A fundamental epidemiological parameter measuring the potential spread of infection is represented by the reproduction number *R*, which is defined as the number of secondary cases generated by a typical infector in a partially immune population. In our simulations, we explore scenarios of *R* ranging from1.3 to 1.9, therefore encompassing estimates of *R* associated to SARS-CoV-2 transmission dynamics observed in fall of 2020 in Italy.

We use a well-known relation (17) between the reproduction number, the distribution of the generation time, and the exponential epidemic growth rate *r* to estimate the reproduction number in model simulation:

$$R = \frac{\boldsymbol{r}}{\sum_{i=1}^{n} \boldsymbol{y_i}(\boldsymbol{e^{-ra_{i-1}}} - \boldsymbol{e^{-ra_i}})/(\boldsymbol{a_i} - \boldsymbol{a_{i-1}})}$$

where $a_0$, $a_1$,..., $a_n$, are the category bounds of the histogram of the generation time, $y_1$, $y_2$, ..., $y_n$ are the corresponding relative frequencies where the observed generation time are within these bounds, and *r* is the exponential growth rate derived from the analysis of the number of new cases over time in the simulated epidemics.

##### 1.4 Simulated intervention strategies

**Reactive class closure based on syndromic surveillance**

In this study, we explicitly model the case isolation, contacts quarantine, and reactive class-closure policy as implemented in Italy since mid-September 2020. The strategy is based on identification of infections among symptomatic individuals in the population using reverse transcription polymerase chain reaction (RT-PCR) testing. The simulated strategy entails the following steps:

- If an individual shows respiratory symptoms and/or fever, they are tested with RT-PCR with probability $\alpha_p = 0.5$ if they are a non-student population or $\alpha_s = 0.95$ if they are a student. The larger probability of being tested used for students stems from the routine temperature screening adopted in most Italian schools at the time. Other values of $\alpha_s$ and $\alpha_p$ are explored in the sensitivity analyses.
- While waiting for sample collection ($T_{ST}$ days after symptom onset, see Tab. S1) and test result ($T_{TR}$ days after sample collection, see Tab. S1), the symptomatic individual is precautionary quarantined in their place of residence, while the other members of their households are allowed to continue their normal activities.
- If the result of the test is negative, the tested individual goes back to their normal activities.
- If the result of the test is positive, then:
    - If they are a student, teaching activities for their class are suspended (while the other classes of their school remain open). Note that although their class is closed and thus their classmates cannot attend school, they are not quarantined and thus

could potentially infect their household members and other individuals in the general population (should they be infectious).

- Regardless of whether the positive individual is a student or not, they are isolated at home for 14 days starting with the date of laboratory confirmation.
- Considering a compliance rate q=95%, the household members of a positive individual are tested with RT-PCR and are quarantined at home for 2 weeks starting from the date of laboratory confirmation.
  - If any of their household members is confirmed to be positive with RT-PCR:
    - They are isolated for 14 days (starting from the date of laboratory confirmation).
    - Moreover, if they are a student, the teaching activities in their class are suspended (starting from the date of confirmation), while the other classes in their school remain open. Note that although their class is closed and thus their classmates cannot attend school, they are not quarantined and thus could potentially infect their household members and other individuals in the general population (should they be infectious).

It is important to stress that other (not individually targeted) social distancing measures were implemented in Italy (e.g., closure of gyms, limited size of gatherings, use of masks). All those interventions are taken into account in the model as an ensemble, by considering different values of the reproduction number (as estimated in the absence of the test-based interventions mentioned above).

**Reactive class closure based on rapid antigen screening**

Antigen-based tests are commonly used in the diagnosis of respiratory pathogens, including influenza viruses and respiratory syncytial virus. The U.S. Food and Drug Administration (FDA) has granted emergency use authorization (EUA) for antigen tests to identify SARS-CoV-2 (18). Antigen tests are immunoassays that detect the presence of a specific viral antigen, which implies current viral infection. Most of the currently authorized tests can be used at the point of care, return results in approximately 15 minutes, and are relatively inexpensive compared to RT-PCR tests. Antigen tests for SARS-CoV-2 are generally less sensitive than RT-PCR for detecting the presence of viral nucleic acid.

To estimate the sensitivity and specificity of antigen tests for SARS-CoV-2 through a meta-analysis, we conducted a literature review in PubMed and Web of Science. Seven studies reporting the sensitivity and specificity of nine rapid SARS-CoV-2 antigen-detection tests were included: Chaimayo 2020 (19); Diao 2020 (20); Lambert-Niclot 2020 (21); Mertens 2020 (22); Nalumansi 2020 (23); Porte 2020 (24); Weitzel 2020 (25). The study by Weitzel et al. compared three antigen tests. The original sensitivity and specificity estimates extracted from the identified studies are summarized in Tab. S3. We estimated the overall sensitivity and specificity through our meta-analysis. A random-effect model was used to estimate the pooled sensitivity and specificity, which resulted to be 69% (95%CI: 41%-97%) and 99% (95%CI: 97%-100%), respectively (Figure S1). For simplicity, in the model we consider 100% specificity.

**Table S3.** Summary of the original sensitivity and specificity used in the meta-analysis.

| Study | Sensitivity (mean, 95% CI) | Specificity (mean, 95% CI) |
|---|---|---|
| Chaimayo 2020 | 98% (91%, 100%) | 99% (97%, 100%) |
| Diao 2020 | 76% (69%, 81%) | 100% (91%, 100%) |
| Lambert-Niclot 2020 | 50% (40%, 61%) | 100% (92%, 100%) |
| Mertens 2020* | 58% | 100% |
| Nalumansi 2020 | 70 % (60%, 79%) | 92% (87%, 96%) |
| Porte 2020 | 94% (87%, 97%) | 100% (92%, 100%) |
| Weitzel 2020 [A] | 62% (51%, 72%) | 100% (89%, 100%) |
| Weitzel 2020 [B] | 17% (10%, 17%) | 100% (89%, 100%) |
| Weitzel 2020 [C] | 85 % (76%, 91%) | 100% (89%, 100%) |

Note: *95% CI of the sensitivity and specificity is not available in this study.

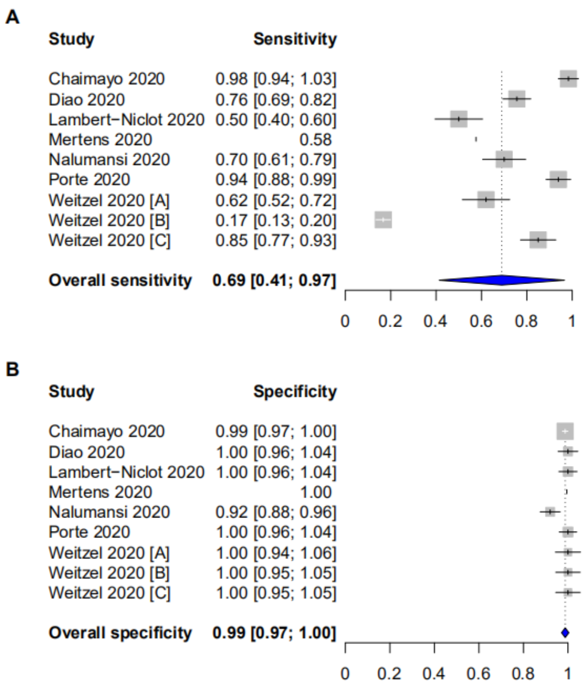


**Fig. S1. Results of the meta-analysis about the sensitivity and specificity of antigen tests. A** Sensitivity of antigen tests. **B** Specificity of antigen tests.

Taking the advantage of the timeliness and cost of antigen tests, we propose a reactive class closure based on the periodic antigen screening on all students irrespectively of their symptoms or clinical signs. Note that the regular testing via PCR of symptomatic individuals in the population is considered to remain in place. According to this strategy, as soon as a student result positive either to a RT-PCR test performed within the syndromic surveillance or to an antigen test regularly applied to all students, the closure of the class of the infected student is imposed, following the same procedure already in place for reactive class closure (see previous section).

In our analysis, we explored three different frequencies for antigen screening: every 3 days, every 7 days, and every 14 days. As rapid antigen tests give very timely results, we assume that laboratory diagnosis from these tests are obtained in the same day of the sample collection.

##### 1.5 Model parametrization

**Initialization.** We initialized the population to reflect the epidemiological conditions characterizing Italy in September 2020. We therefore assume that, at the beginning of our simulations, 5% of the Italian population is immune against SARS-CoV-2 infection. This percentage represents the proportion of the Italian population who experienced the SARS-CoV-2 infection during the first-wave of COVID-19, in spring 2020 (26). The same immunity level was assumed across different age groups. Alternative values of the initial fraction of immune population are explored as sensitivity analysis as well as to simulate an epidemiological situation closer to that of spring 2021. Since we are interested only in assessing the effect of the strategy in suppressing SARS-CoV-2 spread and/or reducing COVID-19 burden, we initialized the simulations with one infectious individual randomly chosen among the susceptible population and no further introductions are considered. A sensitivity analysis was performed to explore how model outcomes changes when considering an initial prevalence of 0.1%, 0.2%, 0.5%, and 1% infected individuals (instead of one single index case). The sensitivity analysis shows little/no impact of the initial number of seeds to the outcome of interest in this study (see Sec. 2.2). Nonetheless, it is important to remark that the number of seeds and the dynamics of the imported cases are expected to play a role should the modeling work aim at reconstructing the observed (transient) dynamics of the epidemic (which is outside the scope of this work).

**Model calibration.** In Italy, after the first COVID-19 case was identified in February 2020, all teaching activities were completely suspended. All schools in the entire country remained closed until September 2020. In order to be consistent with epidemiological evidences characterizing the first epidemic wave in Italy, model parameters shaping the transmission potential of SARS-CoV-2

(Equation 1.1) and the relative contribution of households in the transmission of the infection were assumed in such a way to reproduce a reproduction number (*R*) of 1.1 (8) and a household second attack rate (hSAR) of 51.5% (10). In our simulation, we consider that after the school reopening, *R* increases to values in the range of 1.3 to 1.9, as estimated in Italian regions (8).

As direct quantitative estimates of the contribution of school (or school-related) activities to the increase in the overall transmissibility are unavailable, we consider three scenarios. In the first one we assumed that the increased transmission observed in Italy during September can be entirely ascribed to transmission in schools (F100). The other two scenarios (F50 and F25) account for the increase in the number of contacts in the general community related to the reactivation of teaching activities, such as contacts made on transportation means, extracurricular activities, etc.

In scenario F100, we kept the transmission rates in household and the community as estimated for the summer period and set the transmission in school to obtain the target value of the reproduction number, which corresponds to attributing 100% of the observed increase of the reproduction number in September/October to school transmission. We run the calibrated model and estimated the fraction of infections generated in schools, which we denote as $F_S$. In scenarios F50 and F25, we assume that the fraction of infections is 0.5*$F_S$ and 0.25*$F_S$, respectively. To do so, we fixed the transmission rate in household as in F100, and we re-estimate the transmission rates in school and in the community to obtain the target value of the reproduction number and of the fraction of infections occurring in schools. The total school contribution to infections estimated by the model for the three scenarios (F25, F50, and F100) and different values of the reproduction number is shown in Fig. S2.

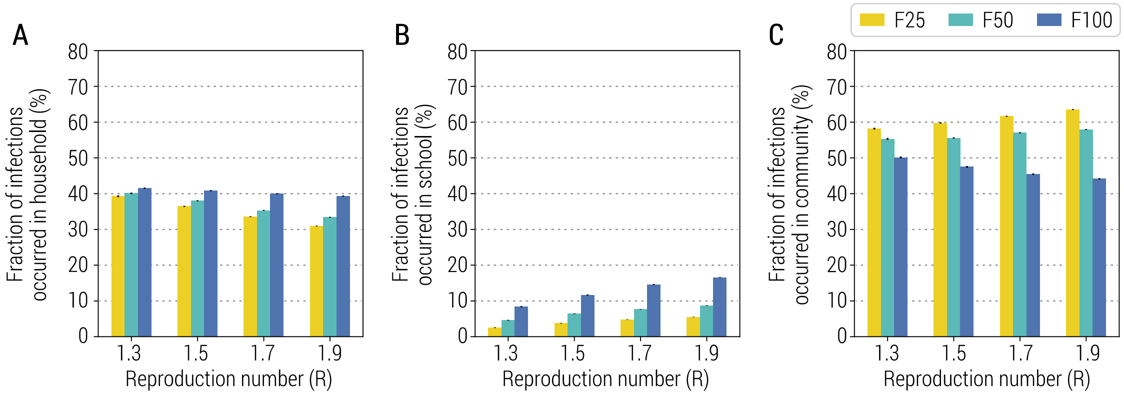


**Fig. S2. Estimated fraction of infections by setting. A** Fraction of infections linked to household transmission for different values of *R* and in the three school transmission contribution scenarios. **B** As A, but for school. **C** As A, but for Community.

#### 2 Reactive class-closure strategy based on syndromic surveillance

##### 2.1 Additional results for the baseline analysis

In addition to the primary results related to COVID-19-related burden reported in in Fig. 1A and 1B, here we show other metrics (Fig. S3). In particular, we estimate that the relative change in the number of symptomatic infections, hospitalized patients, and patients requiring admission to an ICU are similar to those estimated for infections and deaths.

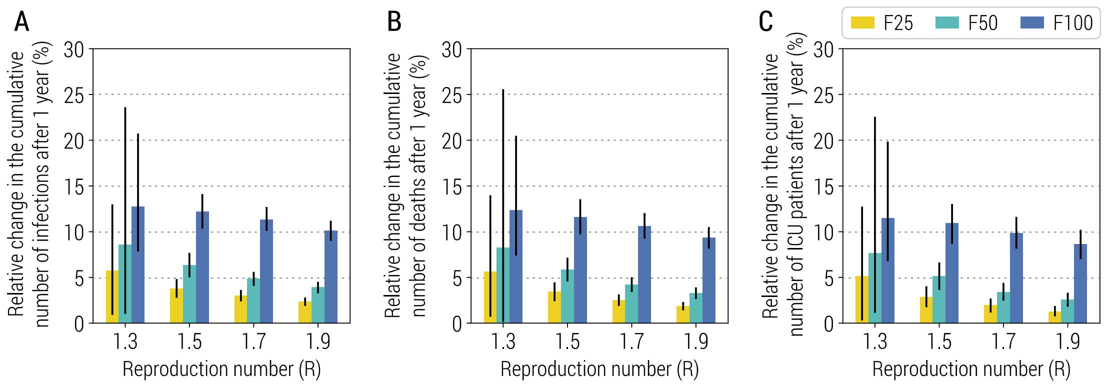


**Fig. S3. Impact of the reactive class-closure policy based on syndromic surveillance. A** Relative change in the cumulative number of symptomatic infections after one year as a function of the reproduction number and for different scenarios about school transmission contribution. The bars correspond to the mean value, while the vertical lines represent 95% quantile intervals; colors refer to the three scenarios F25, F50, F100. Parameters are as the baseline values reported in Tab. S1 and S2. Note that, to exclude spontaneous extinctions from the analysis, only simulations leading to a final infection attack rate of 5% or higher after 1 simulated year are considered. **B** As A, but for the number of hospitalized patients. **C** As A, but for the number of ICU admissions.

In the main text, we showed the results of four sensitivity analyses considering changes in parameters regulating the implementation of the reactive class-closure strategy. In particular, we varied (i) the probability to test a symptomatic student at school, (ii) the probability to test a symptomatic (non-student) individual in the community, (iii) the time from symptom onset to sample collection, and (iv) the time from sample collection to laboratory diagnosis. Fig. S4 shows the impact of these parameters on the number of infections in the student population at the time when the class closure is triggered. For all scenarios, we estimate the number of infected students to be always larger than 10 at the time of class closure, meaning that even a quicker or more intense syndromic surveillance would not be able to readily identify enough infected students to prevent widespread school transmission before classes are closed.

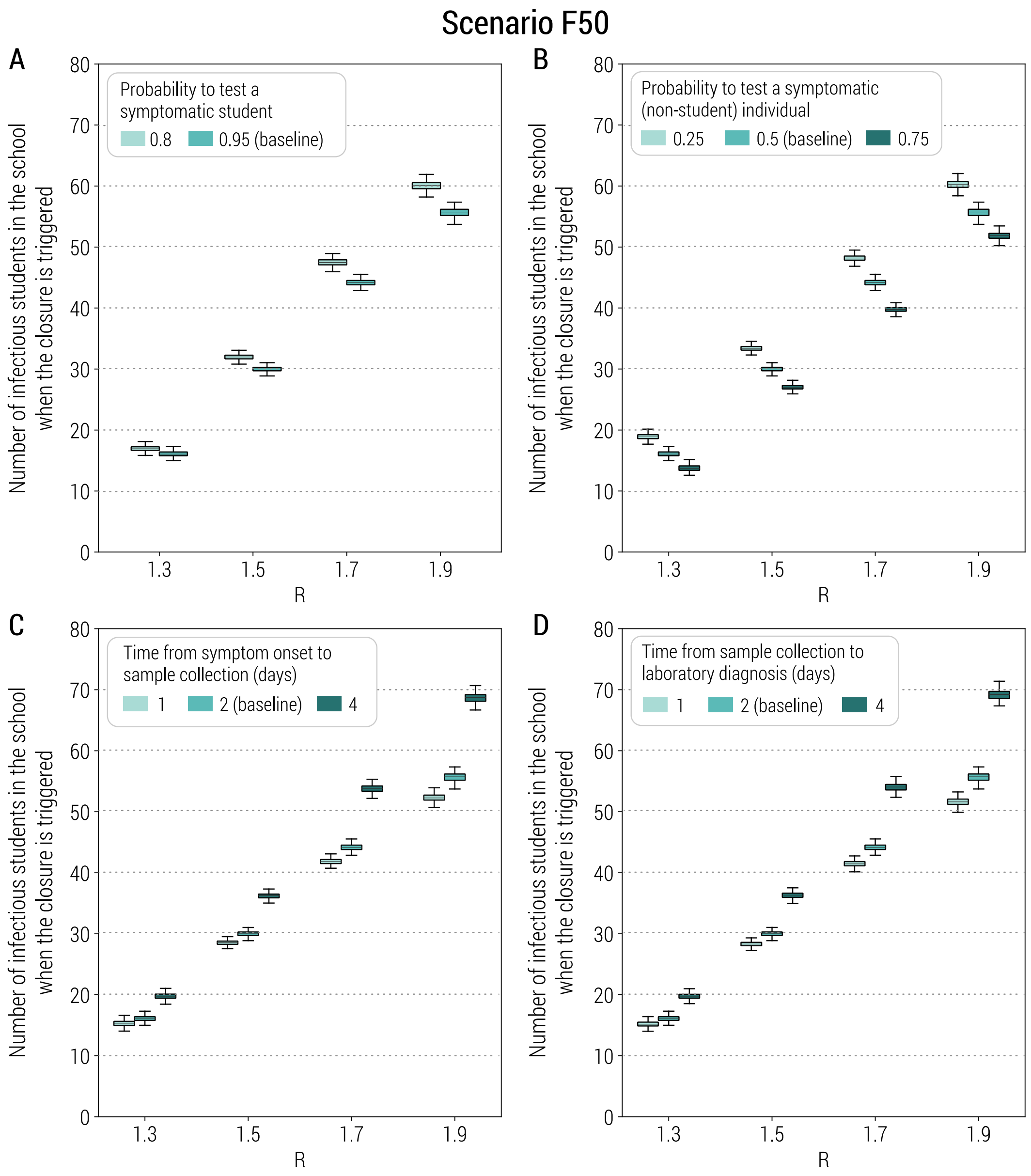


**Fig. S4. Sensitivity of the class-closure strategy based on syndromic surveillance to changes in parameters regulating its implementation. A** Number of infectious students in a school at the time when the class closure is triggered as a function of the reproduction number and for different values of the probability to test a symptomatic student at school. In the boxplot, the middle line corresponds to the median, the lower and upper hinges correspond to the first and third quartiles, the upper whisker extends from the hinge to the largest value no further than 1.5IQR from the hinge (where IQR is the inter-quartile range) and the lower whisker extends from the hinge to the smallest value at most 1.5IQR of the hinge. The same definition of the boxplot is used throughout the manuscript. Parameters are as the baseline and explored values reported in Tab. S1. Note also that *R* is estimated in the absence of the class-closure strategy and the scenario considered is F50. Additionally, to exclude spontaneous extinctions from the analysis, only simulations leading to a final infection attack rate of 5% or higher after 1 simulated year are considered. **B** As A, but for the probability to test a symptomatic (non-student) individual in the community. **C** As A, but for the time from symptom onset to sample collection. **D** As A, but for the time from sample collection to laboratory diagnosis.

##### 2.2 Initial number of seeds

In the baseline analysis, we use 1 seed to initialize the epidemic. To explore the robustness of the results to this choice, we varied the initial number of seeds by assuming four scenarios: 0.1%, 0.2%, 0.5%, and 1.0% of population is considered to be infectious at the start of the simulation. We simulate the impact of the reactive class-closure strategy and found very consistent estimates of the relative change in COVID-19 burden in these 5 scenarios (Fig. S5).

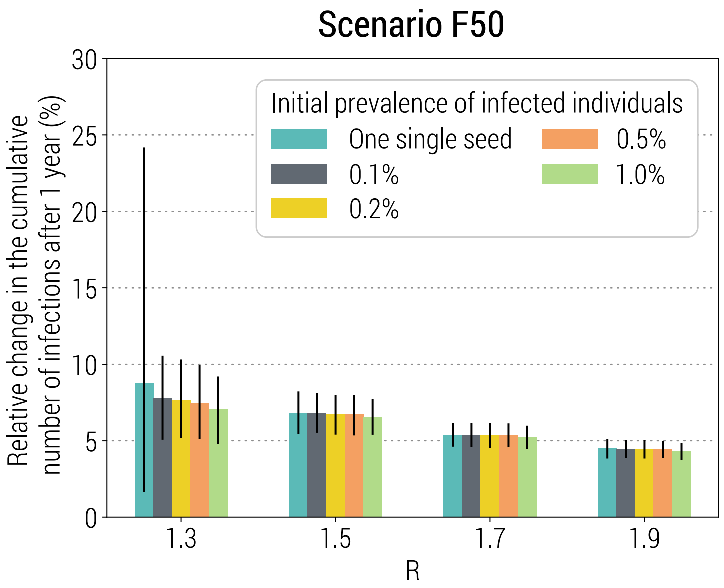


**Fig. S5. Sensitivity of the class-closure strategy based on syndromic surveillance to changes in initial number of seeds used to initialize the epidemic.** Relative change in the cumulative number of infections after one year as a function of the reproduction number and for different initial number of seeds. The bars correspond to the mean value, while the vertical lines represent 95% quantile intervals; colors refer to the baseline value (i.e., 1 seed) and four explored values (i.e., 0.1%, 0.2%, 0.5%, and 1.0% of the simulated population). Note that *R* is estimated in the absence of the class-closure strategy and the scenario considered is F50. Additionally, to exclude spontaneous extinctions from the analysis, only simulations leading to a final infection attack rate of 5% or higher after 1 simulated year are considered.

##### 2.3 Homogeneous susceptibility to infection by age

In the main analysis, we considered age-specific susceptibility to infection (as estimated in the literature (3, 27, 28)). Here we proposed additional sensitivity analyses where we assume a homogenous susceptibility to infection by age (i.e., $\delta_a$=1 for all ages). All the obtained results are very consistent with those obtained in the main analysis (Fig. S6).

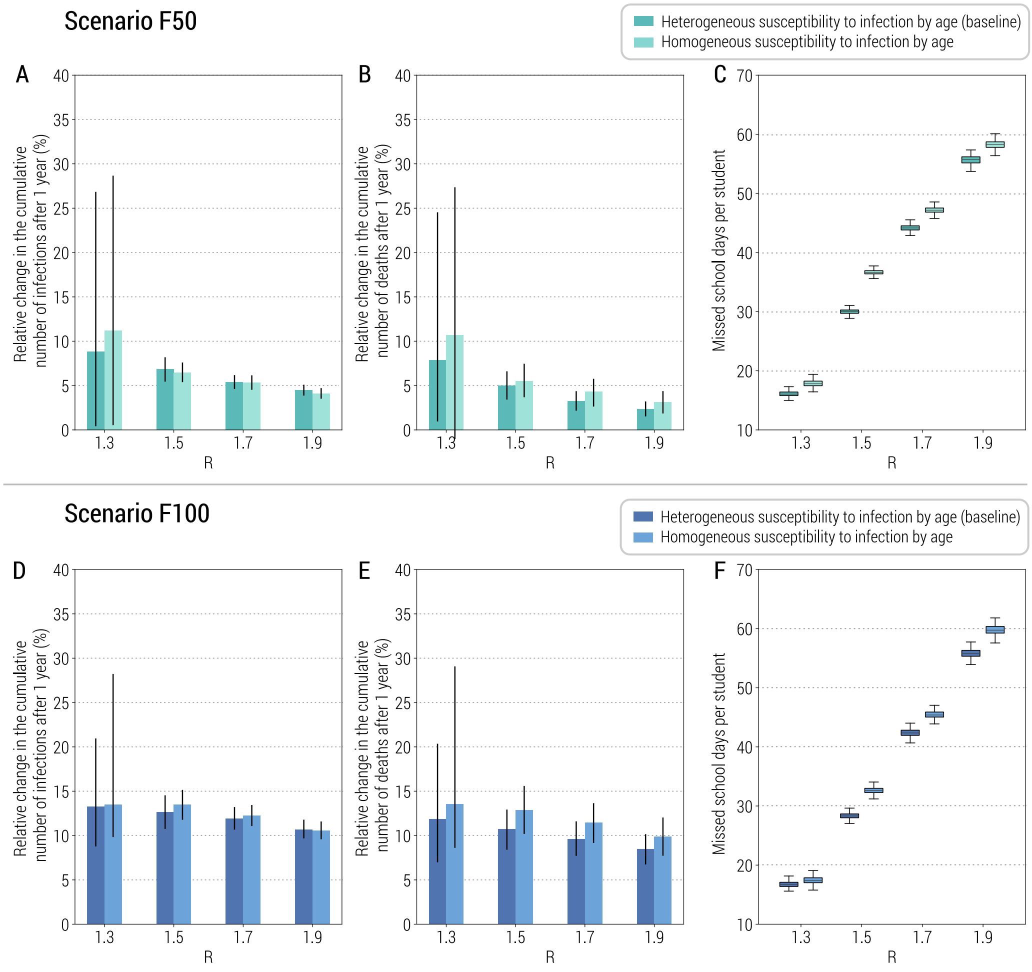


**Fig. S6. Sensitivity of the class-closure strategy based on syndromic surveillance to changes in susceptibility to infection by age. A** Relative change in the cumulative number of infections after one year as a function of the reproduction number and for two scenarios about susceptibility to infection. The bars correspond to the mean value, while the vertical lines represent 95% quantile intervals; colors refer to the age-specific susceptibility (i.e., baseline) and homogeneous susceptibility to infection by age. Note that *R* is estimated in the absence of the class-closure strategy and the scenario considered is F100. Additionally, to exclude spontaneous extinctions from the analysis, only simulations leading to a final infection attack rate of 5% or higher after 1 simulated year are considered. **B** As A, but for the number of deaths. **C** Number of missed school days per student due to the reactive class-closure strategy. **D** As A, but for scenario F50. **E** As B, but for scenario F50. **F** As C, but for scenario F50.

#### 2.4 Infectiousness of asymptomatic vs. symptomatic individuals

In the main analysis, we considered the infectiousness of asymptomatic infected individuals relative to symptomatic one to be equal to 1 (as estimated in the literature (3)). Here we proposed a sensitivity analysis on infectiousness, where we assume that the infectiousness of asymptomatic infected individuals transmit ½ of symptomatic ones. All the obtained results are very consistent with those obtained in the main analysis (Fig. S7).

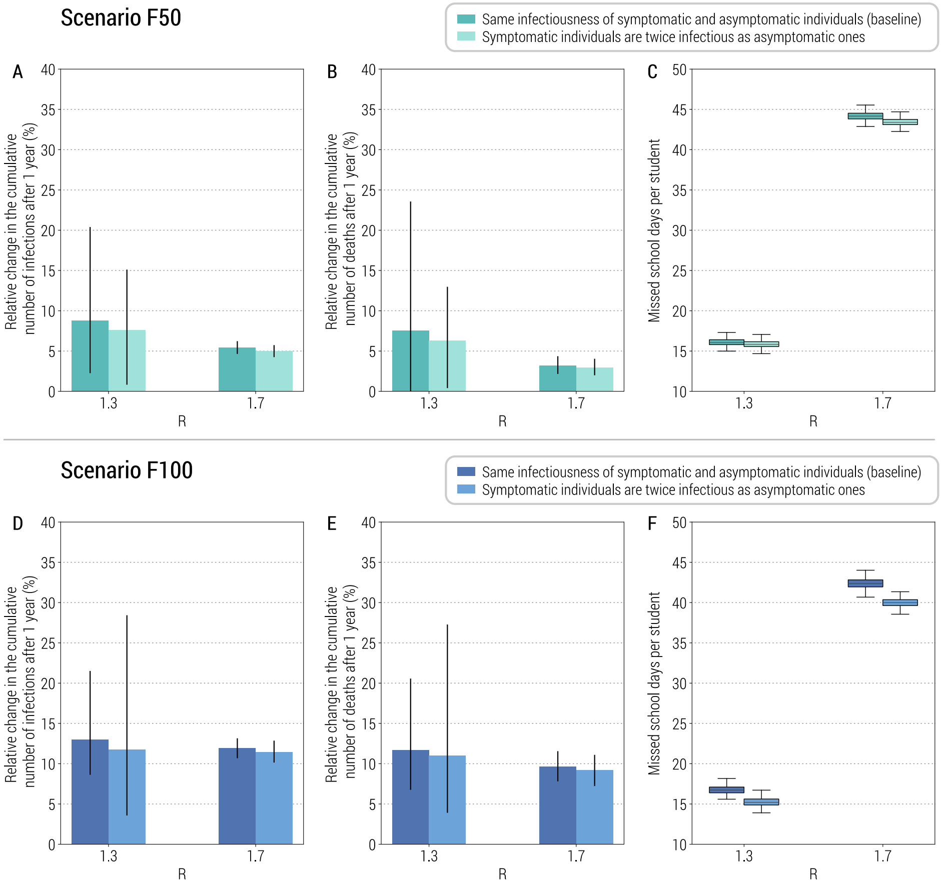


**Fig. S7. Sensitivity of the class-closure strategy based on syndromic surveillance to changes in infectiousness of asymptomatic vs. symptomatic individuals. A** Relative change in the cumulative number of infections after one year as a function of the reproduction number and for two scenarios about relative infectiousness. The bars correspond to the mean value, while the vertical lines represent 95% quantile intervals; colors refer to two assumptions where the same infectiousness between asymptomatic and symptomatic individuals is assumed (i.e., baseline) and asymptomatic individuals are assumed to be 50% less infectious compare to symptomatic cases, respectively. Note that *R* is estimated in the absence of the class-closure strategy and the scenario considered is F100. Additionally, to exclude spontaneous extinctions from the analysis, only simulations leading to a final infection attack rate of 5% or higher after 1 simulated year are considered. **B** As A, but for the number of deaths. **C** Number of missed school days per student due to the reactive class-closure strategy. **D** As A, but for scenario F50. **E** As B, but for scenario F50. **F** As C, but for scenario F50.

##### 2.5 Initial fraction of immune population

In the baseline analysis, we consider 5% of the population to be immune at the beginning of the simulation, according to modeling estimates for the Italian population in September 2020 (26). We performed a sensitivity analysis on the initial immunity of the population by increasing this fraction to 10% and 20%. The obtained results show that the relative change with respect to the simulations without school closure is not affected much by the initial fraction of immune population (Fig. S8 A, B, D, and E). However, with the infection attack rate decreasing as the initial fraction of immune population increases, the number of missed school days due to the class-closure strategy remarkably decreases (Fig. S8 C and F).

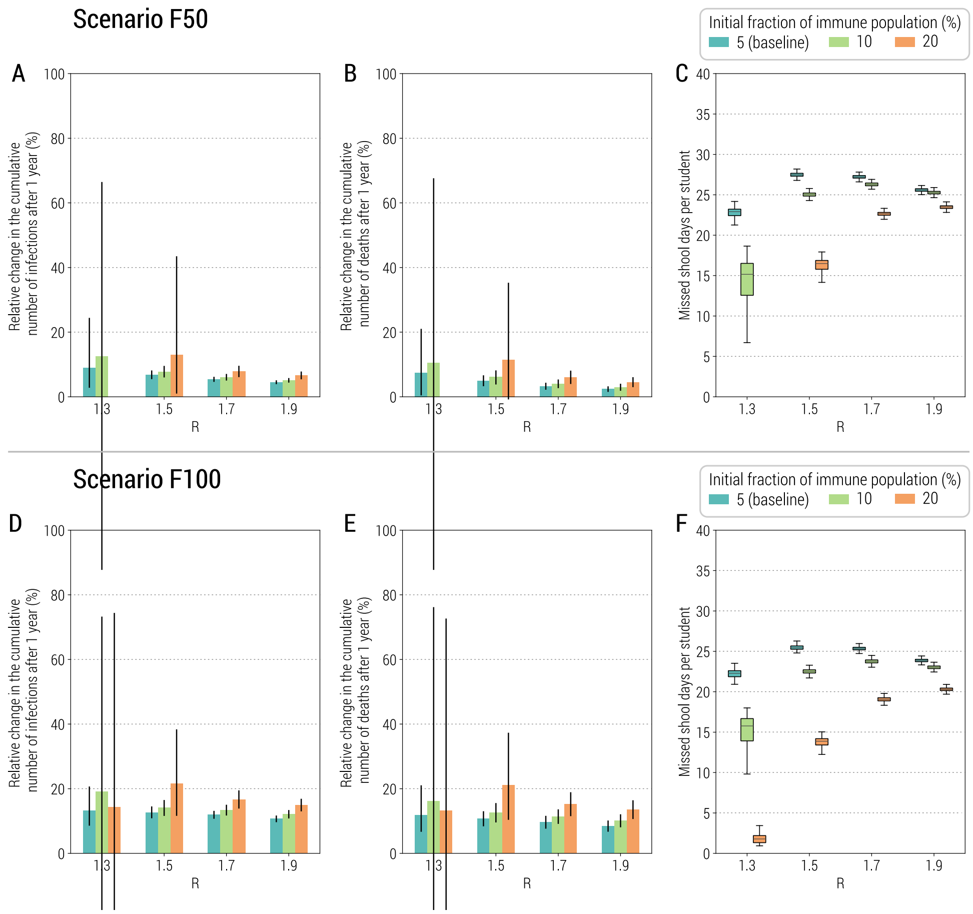


**Fig. S8. Sensitivity of the class-closure strategy based on syndromic surveillance to changes in initial fraction of immune population**. **A** Relative change in the cumulative number of infections after one year as a function of the reproduction number and for two scenarios about relative infectiousness. The bars correspond to the mean value, while the vertical lines represent 95% quantile intervals; colors refer to three assumptions of the initial fraction of immune population, including 5% (i.e., the baseline), 10%, and 20%. Note that *R* is estimated in the absence of the class-closure strategy and the scenario considered is F50. Note also that for *R*=1.3 and F50, the relative change cannot be computed as no simulation without school closure leads to an outbreak when the initial fraction of immune population is 20%. Additionally, to exclude spontaneous extinctions from the analysis, only simulations leading to a final infection attack rate of 5% or higher after 1 simulated year are considered. **B** As A, but for the number of deaths. **C** Number of missed school days per student due to the reactive class-closure strategy. **D** As A, but for scenario F100. **E** As B, but for scenario F100. **F** As C, but for scenario F100.

#### 3 Reactive school-closure strategy based on syndromic surveillance

We implemented and tested a reactive school-closure strategy that mirrors exactly the reactive class closure strategy used in the main analysis; the only difference is that once a closure is triggered, the entire school is closed (instead of the single class where a PCR positive student is confirmed).

Fig. S9 shows the impact of reactive school-closure policy on the COVID-19 burden and the number of missed school days per student due to the strategy. In the baseline analysis, 10% of initially immune population is used. As compared to the class-closure strategy (Fig. 1 of the main text), the school-closure strategy leads to a remarkably higher reduction of COVID-19 burden (Fig. S9 A and B). However, this strategy entails more than 100 missed school days per student (i.e., half of the school year) in most cases (Fig. S9C).

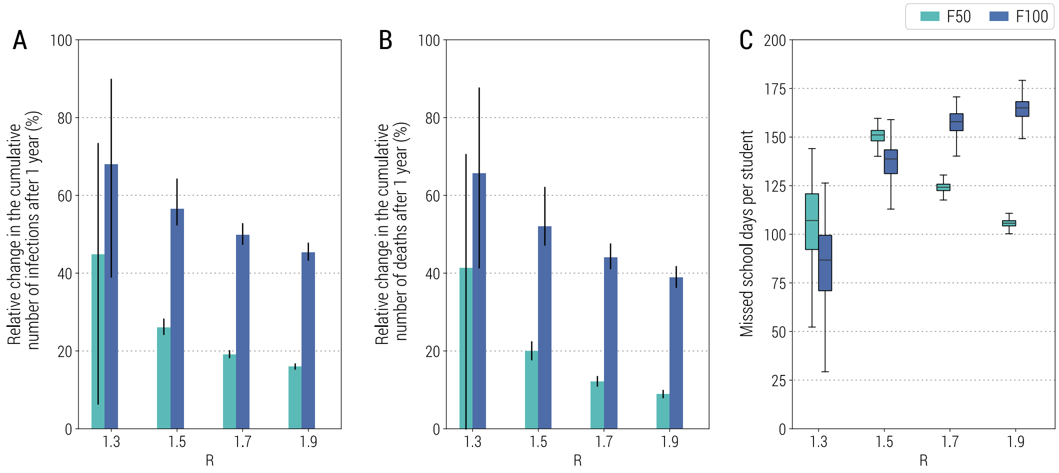


**Fig. S9. Impact of the reactive school-closure strategy based on syndromic surveillance. A** Relative change in the cumulative number of infections after one year as a function of the reproduction number and for different scenarios about school transmission contribution. The bars correspond to the mean value, while the vertical lines represent 95% quantile intervals; colors refer to the two scenarios F50 and F100. Parameters are as the baseline values reported in Tab. S1 and S2. Note that $R$ is estimated in the absence of the class-closure strategy. The relative change is defined as the estimated number of infections after 1 year since the introduction of the first infected individual without the implementation of the school-closure strategy minus the one with the school-closure strategy implemented, relative to the estimated number without the implementation of the school-closure strategy. Note that, to exclude spontaneous extinctions from the analysis, only simulations leading to a final infection attack rate of 5% or higher after 1 simulated year are considered. **B** As A, but for the number of deaths. **C** Number of missed school days per student due to the reactive school-closure strategy.

We performed a sensitivity analysis on the initial immunity of the population by increasing the fraction of immune population to 15% and 20%. Fig. S10 shows the impact of increased fraction of immune population on the COVID-19 burden and the number of missed school days per student due to the reactive school-closure strategy. As observed for the reactive class-closure strategy (Fig. S8), the reduction of COVID-19 burden is not much affected by the initial fraction of immune population (Fig. S10 A and B). However, the number of missed school days decreases with increases in immunity for low values of *R*, while it increases for *R*=1.7 and 1.9 (Fig. S10 C).

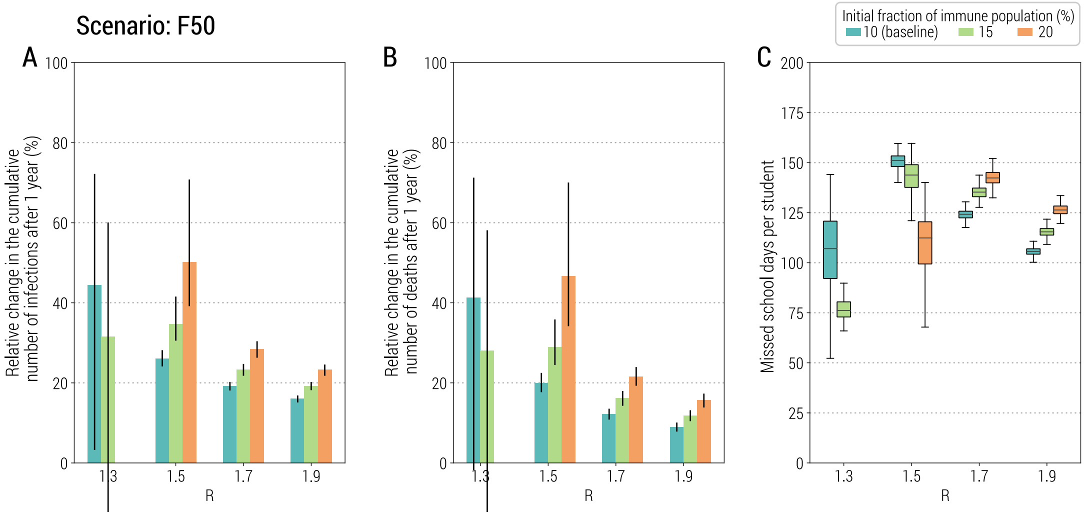


**Fig. S10. Sensitivity of the school-closure strategy based on syndromic surveillance to changes in initial fraction of immune population**. **A** Relative change in the cumulative number of infections after one year as a function of the reproduction number and for three different initial fractions of immune population. The bars correspond to the mean value, while the vertical lines represent 95% quantile intervals; colors refer to three assumptions of the initial fraction of immune population, including 10% (i.e., baseline), 15%, and 20%. Note that *R* is estimated in the absence of the class-closure strategy and the scenario considered is F50. Additionally, to exclude spontaneous extinctions from the analysis, only simulations leading to a final infection attack rate of 5% or higher after 1 simulated year are considered. **B** As A, but for the number of deaths. **C** Number of missed school days per student due to the reactive school-closure strategy.

### 4 Reactive class-closure strategy based on rapid antigen screening

#### 4.1 Additional results for the baseline analysis

In addition to the primary results of COVID-19-related burden shown in Fig. 4A of the main text, Fig. S11 shows more metrics related to the burden of COVID-19, including number of symptomatic infections, number of hospitalized patients, number of patients admitted to ICU, and number of deaths. The results highlight that antigen-based class-closure strategy is capable to prevent not only a big share of infections, but also a significant share of hospitalizations, ICU use, and deaths (Fig. S11).

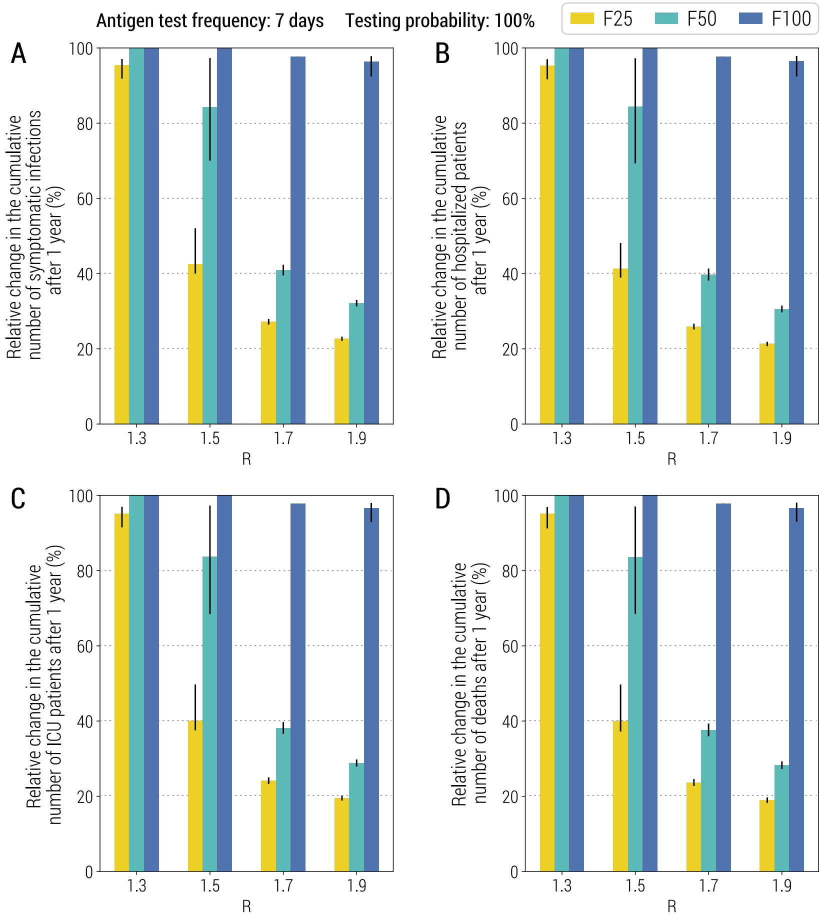


**Fig. S11**. **Impact of the reactive class-closure policy based on antigen screening**. **A** Relative change in the cumulative number of symptomatic infections after one year as a function of the reproduction number and for different scenarios about school transmission contribution. The bar corresponds to the mean value, while the vertical line represents the 95% quantile intervals; colors refer to the three scenarios F25, F50, F100. The fraction of immune population at the beginning of epidemic is set at 10%, the probability of testing a student at school with the antigen test is 100%, the frequency of the antigen screening is weekly; other parameters are as the baseline values reported in Tab. S1 and S2. Note that *R* is estimated in the absence of the class-closure strategy. Additionally, to exclude spontaneous extinctions from the analysis, only simulations leading to a final infection attack rate of 5% or higher after 1 simulated year are considered. **B** As A, but for the number of hospitalized patients. **C** As A, but for the number of ICU admissions. **D** As A, but for the number of deaths.

##### 4.2 Antigen screening frequency

In the main text we explored the impact of antigen screening frequency on the effectiveness of the strategy by performing a sensitivity analysis on screening frequency, where we decreased the screening frequency from once every 3 days to once every 7 days to once every 14 days. Here we show other metrics. Similar to the decrease in the number of infections shown in Fig. 4E (of the main text), the strategy impact on COVID-19 burden (measured through the change in hospitalizations, ICU use, and deaths) tend to decrease with screening becoming less frequent (Fig. S12 A-D). The effectiveness of the control over epidemic decreases mostly due to the low number of infectious students at the time the class closure is triggered as compared to strategy based on syndromic surveillance (Fig. S12E vs. Fig. 2 of the main text).

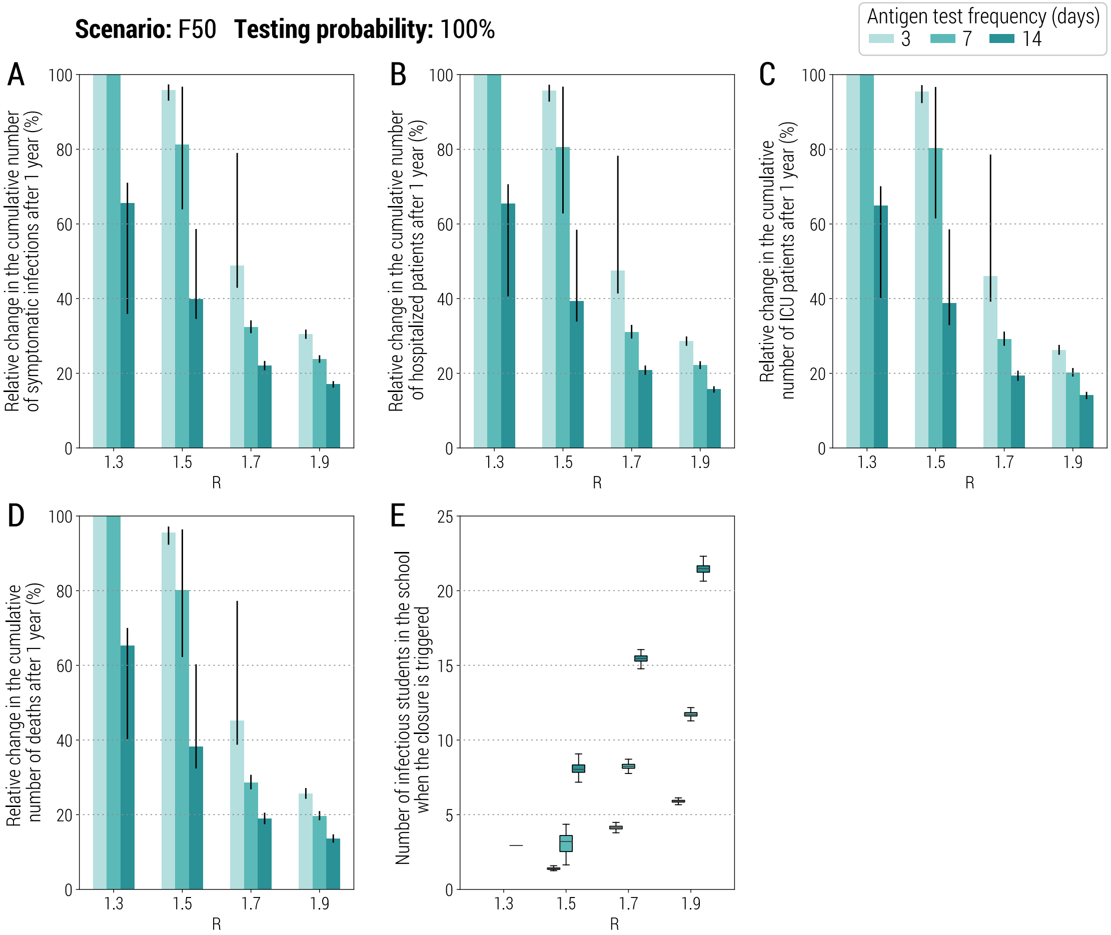


**Fig. S12**. **Sensitivity of the class-closure strategy based on antigen screening to changes in screening frequency. A** Relative change in the cumulative number of symptomatic infections after one year as a function of the reproduction number and for three scenarios about screening frequency (every 3, 7 or 14 days). The bars correspond to the mean value, while the vertical lines represent 95% quantile intervals. Note that *R* is estimated in the absence of the class-closure strategy and the scenario considered is F50. Additionally, to exclude spontaneous extinctions from the analysis, only simulations leading to a final infection attack rate of 5% or higher after 1 simulated year are considered. **B** As A, but for the number of hospitalized patients. **C** As A, but for the number of ICU admissions. **D** As A, but for the number of deaths. **E** Number of missed school days per student due to the reactive class-closure strategy.

In the main text, we also tested the performance of a strategy where, instead of screening all students in one single day once per week, 1/7 of the student population is tested every day. Similar to the results illustrated in Fig. 4F, the impact of the two strategy variations on the COVID-19 burden measured through the change in hospitalizations, ICU use, and deaths is comparable (Fig. S13A-D). When 1/7 of the student population is tested every day, the number of missed school days is higher than the alternative for all values of *R* (Fig. S13E), while the number of infectious students at closure is slightly higher for lower values of *R* and lower for higher values of *R* (Fig. S13F).

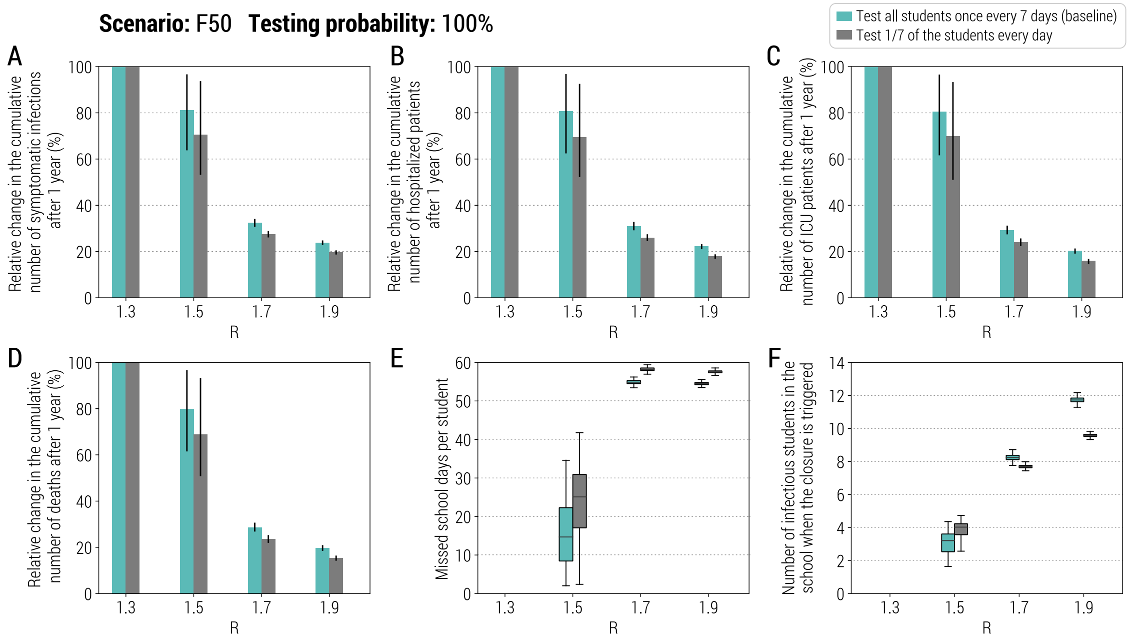


**Fig. S13**. **Sensitivity of the class-closure strategy based on antigen screening to changes in the performance of the strategy. A** Relative change in the cumulative number of symptomatic infections after one year as a function of the reproduction number when all students are tested in one day, once per week, or when 1/7 of the students at each school are tested every day. The bars correspond to the mean value, while the vertical lines represent 95% quantile intervals; colors refer to different performance of the strategy. Note that *R* is estimated in the absence of the class-closure strategy and the scenario considered is F50. Additionally, to exclude spontaneous extinctions from the analysis, only simulations leading to a final infection attack rate of 5% or higher after 1 simulated year are considered. **B** As A, but for the number of hospitalized patients. **C** As A, but for the number of ICU admissions. **D** As A, but for the number of deaths. **E** Number of missed school days per student due to the reactive class-closure strategy. **F** Number of infectious students in a school at the time when the class closure is triggered.

#### 4.3 Initial fraction of immune population

In the analysis of the antigen-based class-closure strategy presented in the main text, we considered 10% of the population to be initially immune. We performed a sensitivity analysis by increasing the fraction of the initially immune population to 15% and 20%. As shown in the main text (Fig. 4H), the strategy performs even better with the higher share of immune population. For 20% of initially immune population, the strategy averts all the hospitalizations, need for ICU and deaths for *R*=1.5, and decreases them by almost 80% for *R*=1.7 (Fig. S14A-D). Significant decrease in the number of infectious students at the moment of class closure illustrates the increasing effectiveness of the strategy (Fig. S14E).

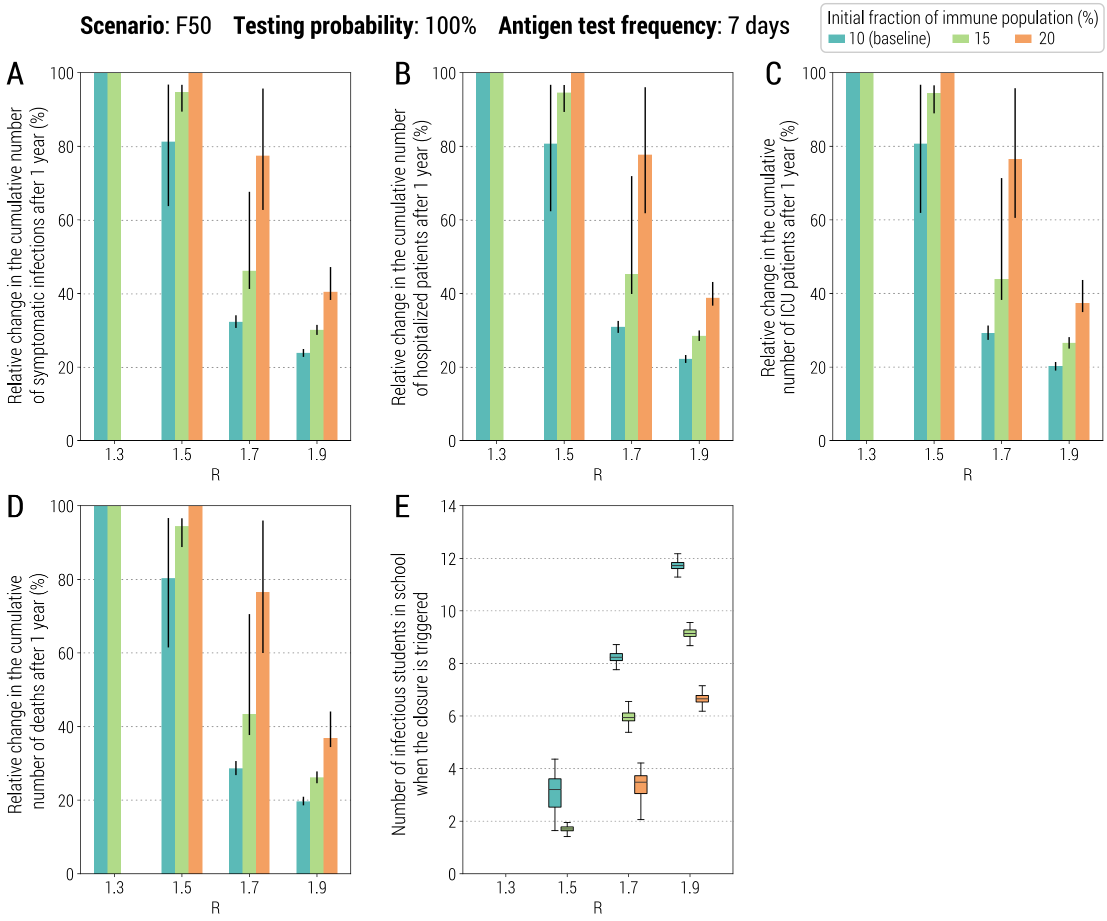


**Fig. S14**. **Sensitivity of the class-closure strategy based on antigen screening to changes in initial fraction of immune population**. **A** Relative change in the cumulative number of symptomatic infections after one year as a function of the reproduction number and for three different initial fractions of immune population. The bars correspond to the mean value, while the vertical lines represent 95% quantile intervals; colors refer to three assumptions of the initial fraction of immune population, including 10% (i.e., baseline), 15%, and 20%. Note that *R* is estimated in the absence of the class-closure strategy and the scenario considered is F50. Additionally, to exclude spontaneous extinctions from the analysis, only simulations leading to a final infection attack rate of 5% or higher after 1 simulated year are considered. **B** As A, but for the number of hospitalized patients. **C** As A, but for the number of ICU admissions. **D** As A, but for the number of deaths. **E** Number of missed school days per student due to the reactive class-closure strategy.
